# Genetic Insight into Birt-Hogg-Dubé syndrome in Indian patients reveals novel mutations in *FLCN*

**DOI:** 10.1101/2021.10.31.21264930

**Authors:** Anindita Ray, Esita Chattopadhyay, Richa Singh, Arnab Bera, Mridul Sarma, Mahavir Munot, Unnati Desai, Sujeet Rajan, Pralhad Prabhudesai, Ashish K. Prakash, Sushmita Roy Chowdhury, Niladri Bhowmick, Raja Dhar, Zarir F. Udwadia, Atin Dey, Subhra Mitra, Jyotsna M Joshi, Saurabh Ghosh, Arindam Maitra, Bidyut Roy

## Abstract

**Background:** Birt-Hogg-Dubé syndrome (BHDS) is a rare monogenic condition mostly associated with germline mutations at *FLCN*. It is characterized by either one or more manifestations of primary spontaneous pneumothorax (PSP), skin fibrofolliculomas and renal carcinoma. Here, we comprehensively studied germline mutations in BHDS patients and asymptomatic members from 15 Indian families.

**Methods:** Targeted amplicon NGS and Sanger sequencing was performed to detect germline mutations at *FLCN* in 31 clinically diagnosed patients and 74 asymptomatic family members. Functional effects and protein-protein interaction of *FLCN* variants were evaluated *in-silico* and molecular docking method. Family-based association study between pathogenic mutations and BHDS was also performed. Germline mutations at genes associated with phenotypically similar diseases were also addressed in few families.

**Results:** Six different types of pathogenic *FLCN* mutations were observed in the patients. Two of them: 11-nucleotide deletion (*c*.*1150_1160delGTCCAGTCAGC*) and splice acceptor mutation (*c*.*1301-1G>A*), were novel mutations. Two unreported Clinvar pathogenic mutations: stop-gain (*c*.*634C>T*) and 4-nucleotide duplication (*c*.*1329_1332dupAGCC*), and known mutations: hotspot mutation (*c*.*1285delC*) and splice donor mutations (*c*.*1300+1G>A*) were also detected. All these mutations greatly affected the protein stability and FLCN-FNIP2 protein interaction. Family-based association studies suggested pathogenic *FLCN* mutations are significantly associated with BHDS. Two pathogenic SNPs, *rs1801133* and *rs138189536*, at *MTHFR*, associated with Homocystinuria, were found in one family.

**Conclusion:** Pathogenic mutations at *FLCN* may play key roles in deregulating metabolic pathways leading to disease pathogenesis. Instead of *FLCN* mutations, *MTHFR* pathogenic SNPs were also detected in clinically diagnosed BHDS patients, therefore, genetic evaluation is necessary to avoid confounding diagnosis.

## Introduction

Birt-Hogg-Dubé syndrome (BHDS) [MIM: 135150]) is a rare inherited condition, first described in 1977 with skin fibrofolliculomas on the forehead, neck and upper torso of patients^1^. Presence of familial primary spontaneous pneumothorax (PSP; [MIM: 173600]) and/or lung cysts, and chromophobe or oncocytic renal cell carcinoma are two other manifestations in BHDS^2,3^. It follows autosomal dominant pattern of inheritance with incomplete penetrance ^4^ and is known to be monogenic, associated with germline mutations at *Folliculin* (*FLCN*; NM_144997.5) located at *17p11*.*2*^5^. More than 290 pathogenic germline mutations at *FLCN* have been reported in BHDS^6^. Among these, a hotspot protein truncating mutation in a hypervariable poly-*C* tract (*C*_8_) in exon 11 (*c*.*1285)* has been reported in different populations^7,8^. BHDS renal tumours exhibit loss of heterozygosity (LOH), suggesting a tumour suppression role of *FLCN* in kidney^9^. Haploinsufficiency at *FLCN* has also been reported in BHDS manifestations^10,11^. FLCN protein is similar to DENN domain proteins, though its exact function is unknown^12^. It interacts via its Carboxy-terminal (C-terminal) with two proteins - Folliculin interacting protein 1 and 2 (FNIP1 and FNIP2)^5^. These complexes play important roles in major metabolic pathways such as modulation of mTOR pathway^13^, AMPK activation^14,15^, PGC1-α regulation, mitochondrial biogenesis^16^, GAP dependant mTORC1 activation^17^, HIFα transcription^18^, cell-cell adhesion^19^, membrane trafficking^20^, autophagy^21^, ciliogenesis^22^, and cell cycle progression^23^.

Skin fibrofolliculomas and pathogenic *FLCN* mutations are two major diagnostic criteria for BHDS, while lung and kidney phenotypes, and presence of first degree family history are known to be minor criteria^24^. However, these manifestations could be population-specific, as skin fibrofolliculomas are not prevalent in East Asian cohorts^25^. A few other similar conditions like Homocystinuria, alpha-1 antitrypsin deficiency, vascular Ehlers-Danlos syndrome, Lymphangioleiomyomatosis (LAMS) may have overlapping pulmonary phenotypes like BHDS, thus confounding disease diagnosis^26^.

Studies of more than 600 BHDS families have been reported world-wide and majority of them from the USA and Europe, fewer from Asia (mostly from East Asia) with only one from India^27^. Here, we have comprehensively profiled germline mutations in BHDS patients and related members from 15 Indian families and predicted molecular mechanisms for disease phenotype.

## Methods

### Ethics Statement

The study was approved by the “Review committee for protection of research risk to humans, Indian Statistical Institute, 2015”. Written informed consent from all adult participants and legal guardians/parents for minors was obtained for the research study using blood samples and subsequent publication of the results.

### Clinical characterization of study population

Patient IDs’ were assigned anonymously for patient families, patients and asymptomatic members such as F1, F2, F3 and so on. We enrolled 31 clinically diagnosed BHDS patients, during 2015 to 2019, with PSP or BHDS-specific lung cysts along with skin and/or renal manifestations, with/without a positive family history and their 74 asymptomatic family members (Supplemental Table S1). This was done with the help of clinicians from different hospitals in India. Clinical phenotype of each patient was also determined using phenotype ontology analysis (HPO) in Phenomizer (Supplemental Methods)^28^.

### Detection of inherited variants at *FLCN* by different sequencing methods

#### Targeted Amplicon Next-generation Sequencing (NGS)

Initially, genomic DNA from blood of 20 patients and 15 related asymptomatic members from 11 families were isolated for targeted amplicon NGS (Supplemental Table S2). Patient F1-1 was included as a positive control, as mutation at *FLCN* was previously determined^27^. All related asymptomatic family-members and patients were not included for NGS study to minimize sequencing manual errors and logistic problems. The 24kb *FLCN* (including UTRs, exons and introns) was amplified by long PCR (Supplemental Table S3a). Exon 6 and its flanking 2.8kb intronic region could not be amplified due to technical limitations but were studied by Sanger sequencing method. Targeted amplicon generation and equimolar pooling were performed (Supplemental methods) before library preparation, which was done using Nextera XT Library Preparation kit (Illumina Inc.). Paired-end 100bp sequencing was performed in Illumina HiSeq 2500 platform. Standard pipelines were followed for data analysis and germline mutations were called by three variant callers such as Haplotype Caller, STRELKA, VarScan2 (Supplemental Methods)^29–35^.

#### Inherited variants in remaining samples and validation of pathogenic variants

Bidirectional Sanger sequencing of all exons (Supplemental Table S3b) was performed for members of 4 more families; F12, F13, F14 and F15 (9 patients and 20 asymptomatic family members), and validation of pathogenic variants (discovered from NGS) in all individuals from 11 families. BioEdit and *in-silico* tools were used for sequence alignment and variant analysis (Supplemental methods)^36,37^.

### e-QTL analysis for expression and population frequencies of *FLCN* variants

Effect of non-coding germline variants on *FLCN* expression (if any) were examined using computed expression quantitative trait loci (e-QTL) data from GTEx^38^. Since BHDS is a rare disease, population-specific alternate allele frequencies of non-coding variants were also checked and obtained from gnomAD (South Asian)^39^ and GenomeAsia100K (Indian)^40^.

### Pedigree disequilibrium test (PDT) for association study

Pathogenic variants and regulatory SNPs at *FLCN* were tested for association with BHDS in families. PDT is based on a test statistic, *T*, which, for a one-tailed test with 5% significance, was considered as significant if values are ≥ 1.64 (Supplemental methods)^41^.

### Homology modelling and molecular docking

The cryo-EM structure of FLCN-FNIP2-Rag-Ragulator complex (pdb code: *6ulg*) was taken as template for modelling wild-type (wt)/mutant FLCN, wt-FNIP2, wt-Ras-related GTP-binding protein A and protein C (RRAGA and RRAGC) monomers using SWISS-MODEL web server. (Supplemental methods). Monomer-models were visualized at Pymol and validated with PROCHECK^42^. Two sets of molecular docking were performed for wild-type and four mutant monomers of *FLCN*, each with i) wt-FNIP2, wt-RRAGA, wt-RRAGC together (4-protein complex), and ii) wt-FNIP2 (2-protein complex). HADDOCK 2.4 web server *Guru Interface* was used for macromolecular docking with default parameters for running the program and subsequent analysis (Supplemental Methods)^43^.

### Copy number variation of *FLCN*

Read count normalization from *FLCN* NGS data of 35 samples was performed by Seqmonk (Supplemental methods) to provide an initial indication of any *FLCN* copy number variation in patients and asymptomatic members in different families. Subsequently, Taqman copy number assays was performed in 4 families - F3, F4, F9, F15 (Supplemental Table S4) and 23 unrelated healthy controls. Taqman Copy number assays for exon 4 (Hs01200751_cn), exon 8 (Hs01889931_cn), exon 13 (Hs01203178_cn) of *FLCN* and *RNase P* (as reference) were performed in real-time PCR instrument. Ct values of both genes were used to determine copy numbers of *FLCN* (Supplemental methods) following statistical analyses in SPSS.

### Germline mutations at other genes associated with similar pulmonary phenotype

Five families (F6, F7, F8, F9 and F10) having 11 patients and 14 related asymptomatic members (Supplemental Table S5a) were taken for evaluation of germline variants in exons of *SERPINA1* (associated with alpha-1 antitrypsin deficiency), and *MTHFR* and *CBS* (associated with Homocystinuria) by Sanger sequencing methods. Patients taken for *FLCN* amplicon NGS study, were also evaluated for germline variants in *COL3A1* (associated with vascular Ehlers-Danlos syndrome), *TSC1* and *TSC2* (associated with LAMS) (Supplemental Methods, Supplemental Table S5b) by targeted amplicon NGS method.

## Results

### Demography and clinical manifestations

Thirty out of 31 clinically diagnosed BHDS patients presented BHDS lung phenotype, with 10 patients also manifesting skin fibrofolliculomas, with/without a positive family history. Patient, F7-49, only presented skin fibrofolliculomas. Three patients also presented renal cancer (Supplemental Table S6). The male to female ratio was 58% and 41.9% in patients and asymptomatic members, respectively. Age ranged from 18 to 87 years and 7 to 88 years in patients and asymptomatic members, respectively. The number of smokers were observed to be more prevalent in patients than asymptomatic members (Table 1).

**Table 1:** Demography and clinical manifestations of patients (n=31) and asymptomatic members (n=74)

| **Patients (n=31)** | **Demography** |
| --- | --- |
| Mean age ± SD, (range in years.) | 49±15.4, (18 - 87) |
| Males | 58% (n=18) |
| Females | 41.9% (n=13) |
| Smoking Habit (>10years) | 22% (n=7) |
| History of Tuberculosis | 16% (n=5) |
| ***BHDS Diagnostic Criteria*** | **Number of patients\*** |
| Patients with lung cysts or PSP | 96% (n=30) |
| Patients with skin fibrofolliculomas | 35% (n=11) |
| Patients with renal cysts/carcinoma | 9.6% (n=3) |
| Patients having 1<sup>st</sup> degree relative with similar respiratory phenotype | 64% (n=20) |
| **Asymptomatic family members** | **n=74** |
| Mean age ± SD, (range in years.) | 36±19.8, (7 - 88) |
| Males | 58% (n=43) |
| Females | 41.8% (n=31) |
| Smoking Habit (>10years) | 1.3% (n=1) |
| History of Tuberculosis | 6.75% (n=5) |
| ***BHDS Diagnostic Criterion (except respiratory or dermal manifestation)*** |  |
| Asymptomatics with renal cysts/carcinoma** | 2.7% (n=2) |

*\*Some of the patients had more than one phenotype*
*\*\*Two individuals from family F7 only presented renal cell carcinoma, but were clinically not evaluated as BHDS, and were third degree relatives of the index patient, therefore they were considered as asymptomatic related members of the index patient.*

### Clinical Characterization of Patients

Clinical histories of the patients were examined by clinicians from different hospitals. Phenotype ontology analysis revealed 17 HPO terms using Phenomizer (Supplemental Figure S1), which assigned PSP and BHDS to 23 of 31 patients, with significant p-values of ≤ 0.05 (Supplemental Figure S2, Supplemental Table S7). Patient ontology for three patients of family F9 did not qualify for PSP or BHDS. Inconclusive results were obtained for patients, F5-26 and F5-28 (family F5), and F15-99 (family F15), however index patients of both families were significantly assigned with PSP.

### Germline mutations at *FLCN*

An average of 7 million reads per sample were obtained from targeted amplicon NGS data (Supplemental Table S8), and after various quality filters, it revealed a total of 412 variants (Supplemental Figure S3). Variants from homo-polymeric regions (>9) were removed to obtain a total of 76 variants. Among these; 4 exonic and 2 splice region mutations were found to be pathogenic. Sanger sequencing of *FLCN* exons validated these 6 pathogenic mutations detected in NGS with 100% concordance. These pathogenic variants were found in 19 of 31 patients and 16 of 74 related asymptomatic members in 10 families (Figure 1 and Table 2). Remaining 70 variants were found in UTRs and introns of *FLCN* (Supplemental Table S9a and Supplemental Table S9b) and among these; 13 and 12 variants were reported as Clinvar ‘benign’ and non-pathogenic, respectively, in literature. Among the 6 pathogenic variants (Figure 1), two were heterozygous novel mutations: a frame-shift deletion of 11 nucleotides, *c*.*1150_1160delGTCCAGTCAGC (c*.*1150_1160del11)* in exon 10 in family F11, and a splice acceptor mutation, *c*.*1301-1G>A* in intron-exon boundary of exon 12 in family F3 were detected. A stop-gain mutation, *c*.*634C>T* in exon 7, and a frame-shift duplication of 4 nucleotides, *c*.*1329_1332dupAGCC* in exon 12, were found in family F5 and F4, respectively, which were Clinvar ‘pathogenic’ heterozygous mutations but unreported in literature yet. The hotspot heterozygous deletion mutation, *c*.*1285delC* in exon 11 was found in 5 families (previously reported F1, and F12, F13, F14, F15). A reported heterozygous splice donor mutation, *c*.*1300+1G>A* was found in exon-intron boundaries of exon 11 in family F2 (Supplemental Figures S4-1 to S4-6).

**Table 2:** Pathogenic Variants at *FLCN* in 19 patients and 16 related asymptomatic members.

| COORD | SNP ID | HGVS | Consequence | Exon | Annotation | Ref | Alt | Patients | Asymptomatic members | Reports |
| --- | --- | --- | --- | --- | --- | --- | --- | --- | --- | --- |
| 17222646 | rs558699420 | *c.634C>T* | *p.Gln212Ter* | 7 | stop-gain | *C* | *T* | F5-25, F5-26, F5-27 | F5-24, F5-32 | Clinvar pathogenic |
| 17217085-17217095 | - | *c.1150_1160del GTCCAGTCAGC\** | *p.Val384Phefs* | 10 | Deletion | *GTCCAGT CAGC* | *GTCCAGT CAGC/-* | F11-70 | F11-73, F11-74 | novel |
| 17216395 | rs80338682 | *c.1285delC* | *p.His429Thrfs* | 11 | Deletion | *C* | *C/-* | F1-1, F1-2, F12-77, F12-78, F13-82 to F13-85, F14-95, F15-99, F15-101 | F1-4 to F1-7, F12-80, F13-89, F15-105, F15-106 | reported (hotspot) |
| 17215284 | - | *c.1329_1332dup AGCC* | *p.Ala445Serfs* | 12 | Duplication | *AGCC* | *-/AGCC* | F4-18 | none | Clinvar pathogenic |
| 17216379 | rs879255676 | *c.1300+1G>A* | splice donor | 11-12 | splice region | *C* | *T* | F2-9 | F2-10, F2-12 | Reported |
| 17215317 | - | *c.1301-1G>A\** | splice acceptor | 11-12 | splice region | *C* | *T* | F3-13, F3-14 | F3-16, F3-17 | Novel |

*\*novel mutations*
*Abbreviations: COORD: Genomic coordinates, Ref: reference allele, Alt: alternate allele*
**Note:** Pathogenic *FLCN* mutations were detected in 10 out of 15 families recruited in the study. All 19 patients with pathogenic *FLCN* mutations presented PSP and/or BHDS lung cysts. Patient F14-95 harboured all three BHDS manifestations: PSP, skin fibrofolliculomas and renal cysts.
Two more patients (F3-13, F12-77) had skin fibrofolliculomas, and patient F13-82 had renal cell carcinoma.

**Figure 1:**
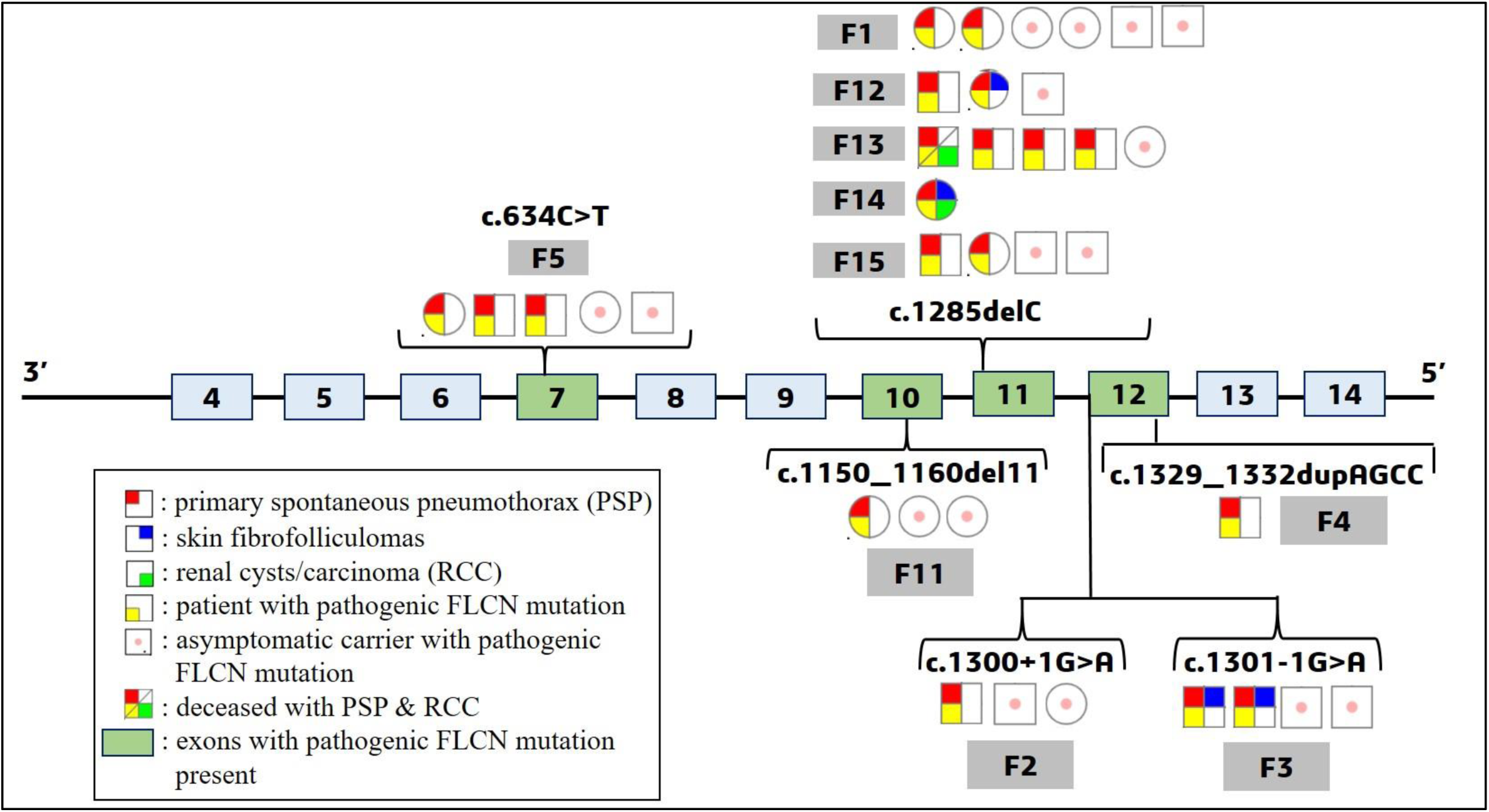
Pathogenic mutations at *FLCN* found in patients and asymptomatic members along with their phenotypes. Legend: Light blue boxes represent exons and green boxes are exons with pathogenic mutations in *FLCN*. Most pathogenic mutations were found between exons 10-13. Circles indicate females, squares males. Family numbers are denoted in grey boxes with patients and asymptomatic members harboring pathogenic *FLCN* mutations. Patient phenotypes are also indicated in red, blue and green colors in individual symbols, and asymptomatic carriers are denoted with a pink dot.

### Expression of *FLCN*: e-QTL study from GTex database

Low *FLCN* expression is reported in tissues (especially in kidney) of BHDS patients^38,44^. Therefore, we checked whether non-coding variants affects *FLCN* expression. We detected 70 non-coding variants and found 12 SNPs (Table 3) common with e-QTL data of *FLCN* expression (for lung and skin tissues - Supplemental Table S10a), with alternate allele frequencies ≤ 15% in South Asian population (Supplemental Table S10b). These SNPs may affect *FLCN* expression since among these; alternate allele genotypes *(CT/TT & AG/GG)* of two SNPs: *rs41345949 (C>T)* and *rs41525346 (A>G)*, were found to be more frequent in patients compared to their asymptomatic family members. The *rs41345949*, although Clinvar benign, is a highly conserved regulatory SNP with an alternate allele frequency of <3% in Indian population. Interestingly, the *TT* and/or *CT* genotypes were found in patients of families without any *FLCN* pathogenic mutations (families F6 and F7). They were also found in patients, F15-99 and F15-101 (genotype *TT*), who also harbour *c*.*1285delC FLCN* mutation, and patient F4-18 (genotype *CT*), also harboring *c*.*1329_1332dupAGCC* mutation. The *rs41525346* is an intronic SNP, highly conserved, and has a distal enhancer-like signature, with an alternate allele frequency of <4% in Indian population.

**Table 3:** Twelve SNPs with median gene expression values in Lung and Skin (exposed) from GTex, and their alternate allele frequencies.

| Sl. No.<br>SNPs | COORD | ID | Annotation | Ref allele | Alt allele frequency<br>(Indian/South Asian) | Lung (median values of gene expression) |  |  |  | Skin-exposed<br>(median values of gene expression) |  |  |  |
| --- | --- | --- | --- | --- | --- | --- | --- | --- | --- | --- | --- | --- | --- |
|  |  |  |  |  |  | REF | HET | ALT | p-value | REF | HET | ALT | p-value |
| SNP 1 | 17236986 | rs41345949 | 5' UTR | C | T= 0.03/0.05 | -0.06(440) | 0.27(69) | N.A. | 1.10E-07 | - | - | - | - |
| SNP 2 | 17212319 | rs7218992 | 3' UTR | C | A= -/0.06 | 0.126(384) | -0.37(119) | -0.91 (12) | 8.90E-14 | 0.05(456) | -0.22(137) | -0.500 (12) | 4.30E-07 |
| SNP 3 | 17213262 | rs12602675 | 3' UTR | C | T= 0.08/0.08 | 0.014(478) | -0.29(37) | N.A. | 0.00011 | 0.02(564) | -0.52(40) | N.A. | 1.10E-09 |
| SNP 4 | 17221501 | rs3744124 | intronic | C | T= 0.11/0.12 | 0.07 (452) | -0.44 (62) | N.A. | 2.40E-09 | 0.06(529) | -0.44(76) | N.A. | 3.40E-18 |
| SNP 5 | 17226931 | rs6502565 | intronic | C | T= 0.12/0.13 | 0.068(451) | -0.44(62) | N.A. | 1.80E-09 | 0.06(528) | -0.46(76) | N.A. | 1.30E-17 |
| SNP 6 | 17226956 | rs76319098 | intronic | C | T= 0.12/0.13 | 0.05(466) | -0.47(47) | N.A. | 1.80E-09 | 0.05(548) | -0.55(56) | N.A. | 1.20E-17 |
| SNP 7 | 17228852 | rs79717038 | intronic | G | A= 0.12/0.13 | 0.06(464) | -0.47(49) | N.A. | 2.70E-08 | 0.06(543) | -0.58(61) | N.A. | 3.10E-19 |
| SNP 8 | 17214314 | rs8067893 | intronic | G | A= -/0.11 | 0.134 (340) | -0.255 (151) | -0.52 (24) | 2.80E-12 | 0.08(407) | -0.23(168) | 0.03 (30) | 1.70E-07 |
| SNP 9 | 17220853 | rs41323249 | intronic | C | T= 0.09/0.11 | 0.14(359) | -0.34 (139) | -0.50(17) | 2.60E-14 | 0.07(430) | -0.12(153) | -0.73 (22) | 4.40E-08 |
| SNP 10 | 17226117 | rs41525346 | intronic | A | G= 0.04/0.08 | 0.08(436) | -0.43(76) | N.A. | 9.90E-08 | 0.06(512) | -0.37(88) | N.A. | 8.70E-07 |
| SNP 11 | 17227704 | rs55836267 | Intronic | T | G= 0.09/0.12 | 0.14(359) | -0.34 (139) | -0.50(17) | 2.60E-14 | 0.07(430) | -0.12(153) | -0.73 (22) | 4.40E-08 |
| SNP 12 | 17221311 | rs41400246 | Intronic | C | T= 0.05/0.10 | 0.13(393) | -0.37 (112) | -0.4 (10) | 2.10E-14 | 0.07(430) | -0.25(129) | -1.11 (13) | 3.00E-10 |

*Abbreviations- COORD: chromosomal co-ordinates, REF: homozygous reference genotype, HET: Heterozygote genotype, ALT: alternate allele genotype, REF: reference allele, ALT: Alternate allele frequency in Indian (GenomeAsia100K) and South Asian (gnomAD) population, N.A.: Gene expression value not available due to none/very low sample size with particular genotype. The numbers in brackets indicate the number of samples harbouring that genotype.*

### Family based test of association between *FLCN* mutations and BHDS by PDT

#### Pathogenic FLCN mutations

Five pathogenic *FLCN* mutations (*c*.*634C>T, c*.*1150_1160del11, c*.*1285delC, c*.*1329_1332dupAGCC, c*.*1300+1G>A*) were tested for family-based association studies. Families F3 and F14 (with *c*.*1301-1G>A* and *c*.*1285delC* mutations, respectively) were not taken as they lacked the required conditions for the test. Calculating *Di* for the 8 families (Supplemental Table S11a), the test statistic (*T*) was found to be 1.83 (≥ 1.64 for significant association). Therefore, the pathogenic *FLCN* variants are significantly associated with BHDS.

#### Regulatory SNPs

Apart from regulatory SNP *rs41345949* observed in eQTL analysis, we observed another *FLCN* SNP, *rs1708629*, affecting *FLCN* expression (Supplemental Table S10a) and previously reported in disease penetrance.^45^ TDT was also performed for these two SNPs for all families (Supplemental Tables S11b and S11c). Calculating for *Di*, where *i* = 1 to 6 (6 families) and *i* = 1 to 5 (5 families) for *rs1708629* and *rs41345949*, respectively; the test statistic (*T*) was found 0.93 for *rs1708629* and 0.85 for *rs41345949*. Therefore, these SNPs were not significantly associated with the BHDS.

### Mutational effects on FLCN protein: *in silico* study

FLCN protein has two distinct terminals: *Longin/N-terminal* (*Lys105* - *Cys265*) and C-terminal (*Pro344 - Met566*) (Supplemental Figure S5a). Three of four frame-shift (*fs*), protein-truncating mutations mapped to the C-terminal: *p*.*Val384Phe*2, p*.*His429Thr*39, p*.*Ala445Ser*11*, while, *p*.*Gln212Ter* mapped to N-terminal (Supplemental Figure S5b) region. All four exonic mutations have pathogenic CADD scores (>15), and result in a premature truncation of the protein (Figure 2, Supplemental Table S12). MaxEnt scores were 8.18 and 8.75 for the splice donor (*c*.*1300+1G>A*) and splice acceptor (*c*.*1301-1G>A*) mutations, and both had pathogenic CADD scores (34 and 33, respectively).

**Figure 2:**
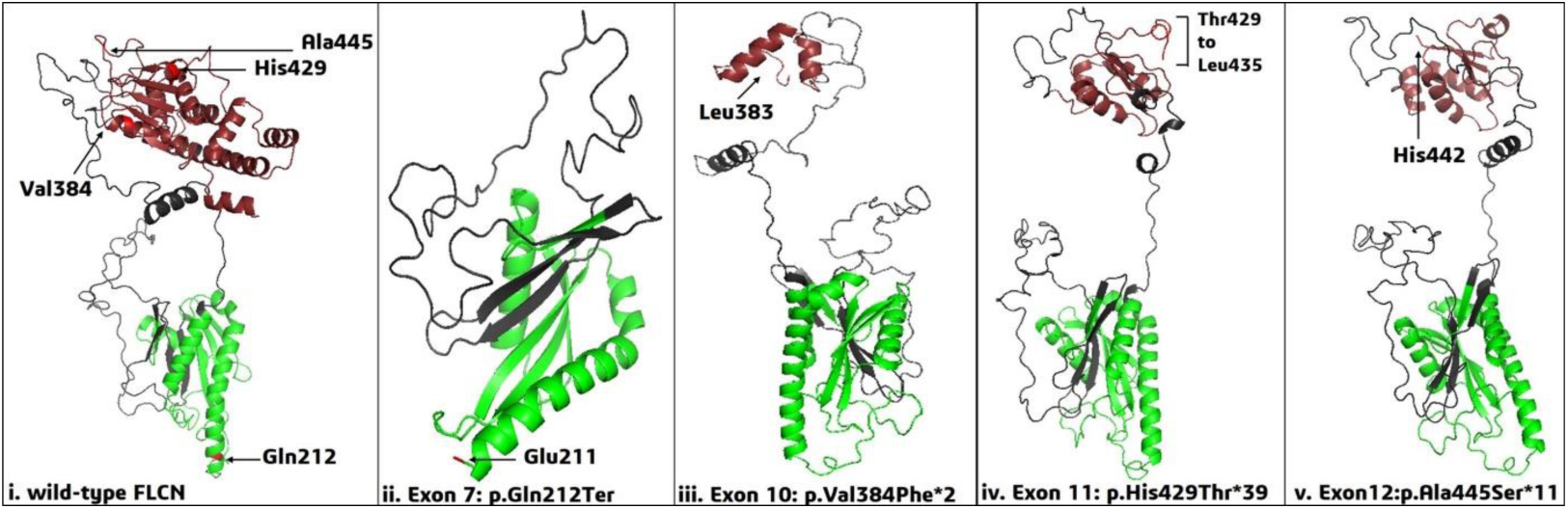
FLCN exonic mutations and their effect on protein structure and interacting proteins. Legend: Green: N-terminal/Longin of FLCN, Brown: C-terminal of FLCN. Structures of homology modelled monomers of FLCN protein – i) wild-type FLCN with amino-acid residues which were affected by mutations; ii) stop-gain mutant FLCN affecting chain termination after Glu211 (i.e., *p*.*Gln212*)*; iii) indel (11-nucleotide) mutant FLCN affecting chain termination after Leu383 (i.e., *p*.*Val384Phe*2)*; iv) indel (single-nucleotide) mutant FLCN, altering the frame from His429 to Thr429, and subsequently creating a stop-codon after Leu435 in the model (i.e., *p*.*His429Thr*39)*; v) four-nucleotide duplicate mutant FLCN, altering the frame and terminating the chain after His442 in the model (i.e., *p*.*Ala445Ser*11)*.

### Homology modelling of wild-type/mutant FLCN and effect on protein-protein interaction

Homology modeled complex of wild type (wt) FLCN or four mutant FLCN (*p*.*Q212*, p*.*V384F*2, p*.*H429T*39, p*.*A445S*11*), with wt-FNIP2, wt-RRAGA, and wt-RRAGC were validated by PROCHECK, with more than 98% residues in the allowed region of the Ramachandran plot (Supplemental Table S13), thus these monomers were accepted for further analyses. Macromolecular docking revealed HADDOCK scores of -342.2±0.0 and -53.1±10.3 for wt-FLCN-FNIP2-RRAGA-RRAGC and wt-FLCN-FNIP2 respectively. This suggested the native FLCN-FNIP2 complex has weaker stability than the wt-4-protein complex. Each of four *FLCN* mutant monomers was docked with either wt-FNIP2-RRAGA-RRAGC or wt-FNIP2 and it resulted in lesser negative HADDOCK scores and low buried surface areas (BSA) than their native complexes (Supplemental Tables S14a & S14b). Less negative HADDOCK scores indicate a lower affinity between interacting partners, and low BSA indicates weaker protein stability. The HADDOCK score of exon-7 mutant-FLCN (*p*.*Q212\**) docked with wt-FNIP2-RRAGA-RRAGC had the least negative score (−203.2±30.5) than the other three mutant FLCN complexes (−252.1±20.6, -264.6±18.1, -242.8±2.8). On the other hand, in two protein docking complex, exon-10 mutant-*FLCN* (*p*.*V384F*2*) had least negative HADDOCK score (−32.0±19.4) followed by exon-11, exon-12 and exon-7 mutant FLCN (−37.6±11.5, -40.2±7.4, and 47.9±11.9, respectively). So, there was a considerable loss of interaction in all mutant complexes (Supplemental Figures S6a and S6b).

### Copy number evaluation of *FLCN* by Taqman assays

NGS data of *FLCN* revealed a difference in log transformed normalized read counts between patients and asymptomatic members particularly in four families (F3, F4, F9 and F10) (Supplemental Figure S7). To validate these differences in normalized read counts, Taqman copy number assay was performed. Ct values were obtained from Taqman assays for exons 4, 8 and 13 of *FLCN*, for all members of the four families and 23 unrelated healthy controls. Normalized Ct values for the exons (dCT or ΔCt) were obtained and transformed to 2^-Δct^ for further analysis (Figure 3). Unpaired t-tests revealed a significant copy number difference for exon 8 in both patients and asymptomatic members compared to unrelated controls (p-values 0.019 and 0.008, respectively). Paired t-tests between patients and asymptomatic members did not reveal any significant copy number difference in any of the three exon assays (exon 4, 8 and 13). But non-parametric test for exon 4 assay and parametric unpaired t-test for exon 13 assay, in patients and asymptomatic members in comparison to unrelated controls did not result any significant copy number difference (Supplemental Table S15).

**Figure 3:**
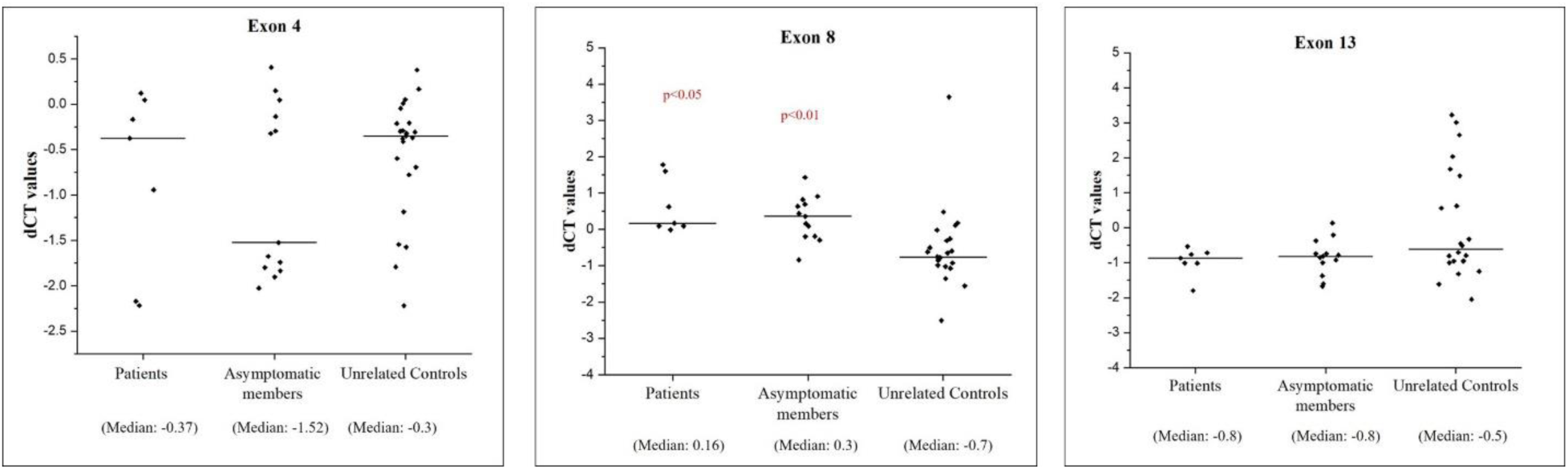
Normalized ΔCt (dCT) values of patients, asymptomatic and unrelated healthy controls obtained from *FLCN* Taqman copy number assays for exons 4, 8 and 13. Legend: For calculation of p-values, 2^-Δct^ values from patients and asymptomatic members were compared with unrelated controls. Patients and unrelated controls (p-value: 0.019), and asymptomatic members and unrelated controls (p-value: 0.008) had a significant difference in copy number only for exon 8. Non-significant p-values have not been shown in the figures.

### Germline mutations at *SERPINA1, MTHFR, CBS, COL3A1, TSC1* and *TSC2*

Absence of *FLCN* pathogenic mutations in 12 patients from 5 families led us to evaluate germline variants in *SERPINA1 MTHFR* and *CBS*, which are known to be associated with other BHDS-like syndrome. Two pathogenic SNPs were found at *MTHFR* - *rs1801133 (C>T)* in exon 5 and *rs138189536 (C>T)* in exon 2, in both patients of family F9 and asymptomatic members, F9-63 and F9-64 (Supplemental Figure S8a and S8b). Both the SNPs are conserved and may affect protein function. Three other SNPs at *SERPINA1*, 5 other synonymous SNPs at *MTHFR* and 2 other synonymous SNPs at *CBS* were found to be Clinvar ‘benign’ (Supplemental Table S16a). Sequencing revealed 32 variants in *COL3A1*, 78 in *TSC1* and 63 in *TSC2*. Among 4 exonic SNPs at *COL3A1*, rs1801183 was detected in only one asymptomatic member F2-10 (Supplemental Table S16b) and found possibly damaging in PolyPhen with a pathogenic CADD score of 23.3. In *TSC1*, among 14 UTR and coding SNPs; one 5’ UTR SNP, rs116951280 (T>A) with a pathogenic CADD score of 16.04, was found in patients F3-13 and F3-14. In *TSC2*, apart from three germline variants (*c*.*482-341G>A, c*.*4990-342G>A, c*.*1120-512C>G*), five synonymous benign SNPs and a non-frameshift heterozygous deletion of 3 nucleotides (*rs1387854239*) were detected in members of family F3.

## Discussion

In this study, BHDS lung phenotype (PSP and/or multiple bilateral lung cysts) was found to be most prevalent followed by skin fibrofolliculomas and renal cysts/carcinoma (Table 1). This observation is in accordance with several East Asian studies, where the lung phenotype is more common (87.3%), than skin lesions (36.7%) and kidney cancer (7.2%), unlike studies from Western countries^25^. These population-specific differences may be due to differential genetic and/or environmental factors contributing to disease pathogenesis.

Twenty-seven of 31 patients was diagnosed with PSP, with recurrent pneumothoraces in 9 patients. Their age of onset ranged from 20 to 59 years. We calculated the probability of recurrence of PSP in 27 patients based on a generalized estimate (GEE) in SPSS. The recurrence of PSP was taken as a dependant variable for ‘age of onset of first spontaneous pneumothorax’, while patient gender, presence/absence of family history, tobacco habits and presence of *FLCN* pathogenic mutations were considered as co-factors (Supplemental Table S17). Analysis revealed a significant association (p-value 0.047) between the age of onset and PSP recurrence. The mean age of patients with recurrent PSP and single PSP in our cohort are 36±13.01 and 40±11.6 years respectively, with age of onset ≤ 25 years in two recurrent PSP patients. A recent study reported that patients with single PSP are significantly older (mean age: 38.9±16) than patients with recurrent PSP (mean age: 29.7±11)^46^. Therefore, age of onset is an important factor for PSP recurrences in patients.

Genotype-specific phenotypes were not observed in this study. Nineteen BHDS patients with pathogenic *FLCN* mutations (Table 2), showed lung phenotype and skin fibrofolliculomas in 4 patients and RCC and renal cysts in 2 patients (Figure 1). One patient with *c*.*1285delC* mutation also presented breast fibroadenoma, which has been reported in another BHDS study with patients negative for pathogenic *FLCN* mutations^47^. Eleven of 19 patients (57.8%) from 5 families harboured known hotspot mutation - *c*.*1285delC*, which is observed in most of the BHDS patients in other studies. Family based association test (PDT) has been sparsely done in BHDS studies. Here, we observed that the novel, unreported Clinvar pathogenic, hotspot and splice donor mutations (Table 2), were significantly associated (Supplemental Table S11a) with BHDS.

Sixteen of 74 asymptomatic members also harbour pathogenic *FLCN* mutations. Mean age, at onset of BHDS phenotype, of 19 patients with pathogenic *FLCN* mutations was 44.1±10.9 yrs, which is much higher than the mean age of 16 asymptomatic members (29.5±20.7 yrs) with *FLCN* mutations. It suggests that few of the asymptomatic members may manifest BHDS after few years. Alternatively, this may be attributed to the presence of 7 minors (aged ≤ 15 years) in the asymptomatic group. All asymptomatic members also need to be clinically evaluated, since we observed an asymptomatic sibling with *c*.*1285del11* mutation harboring several small basal and bilateral lung cysts after clinical re-evaluation in a previous study^27^. Asymptomatic members with pathogenic mutations may harbour un-ruptured pulmonary cysts and abnormal epithelial/mesenchymal interactions in pleura^48^ and may result in PSP later, when combined with other factors.

Homology modelling of interacting proteins with mutant FLCN containing novel, unpublished and hotspot mutant *FLCN* significantly affected the protein structures (Figure 2). Three protein-truncating pathogenic mutations (*p*.*Val384Phefs, p*.*His429Thrfs and p*.*Ala445Serfs*) are present in C-terminal of FLCN which interact with FNIP1/2 and the stop-gain mutation (*p*.*Gln212Ter*) in *longin* domain is necessary for Rag-mediated mTORC1 lysosomal activation. Their protein-interacting docking scores indicated that the FLCN C-terminal mutations substantially reduced the protein stability in the FLCN-FNIP2 complex, while the stop-gain mutation (in longin domain) did so in the 4-protein complex (FLCN-FNIP2-RRAGA-RRAGC).

Large intragenic indels have been reported in BHDS patients^10,49^. Our *FLCN* NGS data showed a read count difference between patients and asymptomatic members of 4 families (F3, F4, F9 and F10). However, Taqman copy number assays for exons 4, 8 and 13 in *FLCN* could not detect a significant difference in copy numbers between patients and asymptomatic members. But a significant copy number difference was only observed when Taqman data for exon 8 in patients and asymptomatics were compared with that of unrelated controls (Figure 3). But we could not validate this difference from NGS data, since we had not taken unrelated controls in the NGS study. Patients of family F9 was later discovered to harbour pathogenic SNPs in *MTHFR*, associated with Homocystinuria. Therefore, on removing family F9 data from Taqman copy number analysis, we found a significant copy number difference in both exons 4 and 8, between patients and unrelated controls (p-value: 0.049 and 0.037, respectively), and between asymptomatic members and unrelated controls (p-value: 0.027) (Supplemental Table S18). Families F3 and F4 harbour splice acceptor (*c*.*1301-1G>A*) and duplication (*c*.*1329_1332dupAGCC*) mutations but family F10 did not harbour any pathogenic *FLCN* mutations. This suggests that there may be other genes that may be associated with disease phenotype and whole-genome sequencing (WGS) may throw light in mutation(s) in, yet not known, other implicated gene/s. These results could show copy number differences between BHDS family members and unrelated controls. More BHDS families need to be studied, since no significant differences were observed for exon 13 assay. Read count results from targeted amplicon NGS send a cautionary note, as their implied read-count differences between patients and asymptomatic members were not detected in subsequent Taqman assay. Therefore, another validation method is necessary for detection of copy number changes.

PSP or BHDS was not detected in the HPO analysis in patients from family F9 (Supplemental Figure S2). Eventually, germline pathogenic SNPs (*rs1801133* and *rs138189536*) were found at *MTHFR* in patients of family F9, who were clinically diagnosed as BHDS. The patients had a positive family history of pulmonary cysts which is a rare presenting feature of Homocystinuria, thus, perhaps, contributing to a misdiagnosis. Similarly, two patients from two different families were initially suspected to have LAMS, but genetic evaluation confirmed those as BHDS patients. Therefore, genetic evaluation is necessary for clinically diagnosed BHDS patients. Pathogenic CADD scores for two SNPs at *COL3A1* and *TSC1* (*rs1801183* and *rs116951280* respectively) were also found in an asymptomatic member, with splice donor *FLCN* mutation and patients with splice acceptor *FLCN* mutation. However, the functional implication of these SNPs in disease pathogenesis could not be ascertained.

### FLCN Mutations and possible mode of molecular pathogenesis

Four protein truncating mutations (Table 2) were detected in this study and it is reported that misfolded *FLCN* proteins, due to truncating mutations, may lead to proteosomal degradation. The hotspot mutation, *c*.*1285dupC (H429Pfs)*, and several C-terminal missense mutations also destabilize the *FLCN*-*FNIP1*/2 binding^50^. BHDS patients in our cohort harbour lung cysts, and it was reported that loss of *FLCN* in murine alveolar cells resulted in dysfunctional activation of AMPK, leading to damaged lung function and apoptotic alveolar cell collapse^15^.

Molecular docking analysis revealed that unpublished stop-gain mutation, p.*Gln212Ter* affects the FLCN-FNIP2-RRAGA-RRAGC binding and stability. It maps to the *longin* domain, where another residue, *p*.*Arg164*, is reported to be an important catalytic residue for GAP activity in mTORC1 activation in lysosomes^17^. Therefore, functional validation to know the role of the stop-gain mutation in this pathway is required. Three protein-truncating mutations were found in the C terminal region of FLCN, which also directly interacts with Rab7A, involved in lysosomal degradation of epidermal growth factor (EGFR). The study in a BHD-RCC cell line resulted in increased cell proliferation, migration and angiogenesis^20^. Therefore, the C terminal region of *FLCN* may have important, yet undiscovered, functions that may involve membrane trafficking in BHDS pulmonary phenotype.

## Conclusion

This is a first comprehensive genetic study from India with 15 BHDS families (31 patients and 74 asymptomatic individuals). We found 10 of 15 families (66.6%) harbour six pathogenic, protein-truncating *FLCN* mutations. Among these: two are novel, two with no published reports, one hotspot and one reported splice donor mutation. They were significantly associated with disease phenotype in family based test (PDT) and found in key functional domains that might greatly affect protein binding and downstream signalling pathways. However, we did not find any pathogenic mutations at *FLCN* in 4 clinically diagnosed and HPO validated BHDS families (F6, F7, F8 and F10). Therefore, we suggest for NGS of *FLCN* whole genome to detect more mutations or whole genome sequencing of these patients to detect other undescribed disease genes. Our findings also suggest for presence of larger mutational spectrum in Indian patients. Therefore, large-scale sequencing studies in more families could truly unveil the genetic landscape of BHDS.

## Data Availability

All data produced in the present study are available upon reasonable request to the authors

## Acknowledgements

Authors would like to thank all patients, their family members, and control healthy individuals for consenting to let their DNA be used in this research. We thank Dr. Ritikha Jha and Dr. Ritabrata Mitra for their clinical insight in the study. Authors are grateful to Mr. Subrata Patra and Mr. Shekhar Ghosh of CoTeRI at NIBMG for their help and support during the NGS experiments. We appreciate and extend heartfelt gratitude to Dr. Analabha Basu, Dr. Roshni Roy, and Ms. Joyeeta Chakraborty for their valuable suggestions during this study.

## Ethics Approval

The study was approved by the “Review committee for protection of research risk to humans, Indian Statistical Institute, 2015”. Written informed consent for the research study and subsequent publication of the results was obtained from all adult participants and legal guardians/parents for minors. All patient data has been anonymized, and any further information may be obtained from corresponding author.

## References

1. Birt AR, Hogg GR, Dubé WJ. Hereditary Multiple Fibrofolliculomas With Trichodiscomas and Acrochordons. Arch Dermatol. 1977. doi:10.1001/archderm.1977.01640120042005

2. Zbar B, Alvord WG, Glenn G, et al. Risk of renal and colonic neoplasms and spontaneous pneumothorax pneurnothorax in the Birt-Hogg-Dubé syndrome. Cancer Epidemiol Biomarkers Prev. 2002.

3. Nickerson ML, Warren MB, Toro JR, et al. Mutations in a novel gene lead to kidney tumors, lung wall defects, and benign tumors of the hair follicle in patients with the Birt-Hogg-Dubé syndrome. Cancer Cell. 2002. doi:10.1016/S1535-6108(02)00104-6

4. Abolnik IZ, Lossos IS, Zlotogora J, Brauer R. On the inheritance of primary spontaneous pneumothorax. Am J Med Genet. 1991. doi:10.1002/ajmg.1320400207

5. Schmidt LS, Linehan WM. FLCN: The causative gene for Birt-Hogg-Dubé syndrome. Gene. 2018. doi:10.1016/j.gene.2017.09.044

6. Kennedy JC, Khabibullin D, Boku Y, Shi W, Henske EP. New Developments in the Pathogenesis of Pulmonary Cysts in Birt-Hogg-Dubé Syndrome. Semin Respir Crit Care Med. 2020. doi:10.1055/s-0040-1708500

7. Schmidt LS, Warren MB, Nickerson ML, et al. Birt-Hogg-Dubé syndrome, a genodermatosis associated with spontaneous pneumothorax and kidney neoplasia, maps to chromosome 17p11.2. Am J Hum Genet. 2001. doi:10.1086/323744

8. Toro JR, Wei MH, Glenn GM, et al. BHD mutations, clinical and molecular genetic investigations of Birt-Hogg-Dubé syndrome: A new series of 50 families and a review of published reports. J Med Genet. 2008. doi:10.1136/jmg.2007.054304

9. Vocke CD, Yang Y, Pavlovich CP, et al. High Frequency of Somatic Frameshift BHD Gene Mutations in Birt-Hogg-Dubé-Associated Renal Tumors. J Natl Cancer Inst. 2005;97(12). doi:10.1093/jnci/dji154

10. Benhammou JN, Vocke CD, Santani A, et al. Identification of intragenic deletions and duplication in the FLCN gene in Birt-Hogg-Dubé syndrome. Genes Chromosom Cancer. 2011. doi:10.1002/gcc.20872

11. Okamoto S, Ebana H, Kurihara M, et al. Folliculin haploinsufficiency causes cellular dysfunction of pleural mesothelial cells. Sci Rep. 2021;11(1):10814. doi:10.1038/s41598-021-90184-9

12. Nahorski MS, Reiman A, Lim DHK, et al. Birt Hogg-Dubé Syndrome-associated FLCN mutations disrupt protein stability. Hum Mutat. 2011. doi:10.1002/humu.21519

13. Chen J, Futami K, Petillo D, et al. Deficiency of FLCN in mouse kidney led to development of polycystic kidneys and renal neoplasia. PLoS One. 2008. doi:10.1371/journal.pone.0003581

14. Baba M, Hong SB, Sharma N, et al. Folliculin encoded by the BHD gene interacts with a binding protein, FNIP1, and AMPK, and is involved in AMPK and mTOR signaling. Proc Natl Acad Sci U S A. 2006. doi:10.1073/pnas.0603781103

15. Goncharova EA, Goncharov DA, James ML, et al. Folliculin Controls Lung Alveolar Enlargement and Epithelial Cell Survival through E-Cadherin, LKB1, and AMPK. Cell Rep. 2014. doi:10.1016/j.celrep.2014.03.025

16. Hasumi H, Baba M, Hasumi Y, et al. Regulation of mitochondrial oxidative metabolism by tumor suppressor FLCN. J Natl Cancer Inst. 2012. doi:10.1093/jnci/djs418

17. Shen K, Rogala KB, Chou HT, Huang RK, Yu Z, Sabatini DM. Cryo-EM Structure of the Human FLCN-FNIP2-Rag-Ragulator Complex. Cell. 2019. doi:10.1016/j.cell.2019.10.036

18. Nishii T, Tanabe M, Tanaka R, et al. Unique mutation, accelerated mTOR signaling and angiogenesis in the pulmonary cysts of Birt-Hogg-Dubé syndrome. Pathol Int. 2013. doi:10.1111/pin.12028

19. Medvetz DA, Khabibullin D, Hariharan V, et al. Folliculin, the product of the Birt-Hogg-Dube tumor suppressor gene, interacts with the adherens junction protein p0071 to regulate cell-cell adhesion. PLoS One. 2012. doi:10.1371/journal.pone.0047842

20. Laviolette LA, Mermoud J, Calvo IA, et al. Negative regulation of EGFR signalling by the human folliculin tumour suppressor protein. Nat Commun. 2017. doi:10.1038/ncomms15866

21. Bastola P, Stratton Y, Kellner E, et al. Folliculin Contributes to VHL Tumor Suppressing Activity in Renal Cancer through Regulation of Autophagy. PLoS One. 2013. doi:10.1371/journal.pone.0070030

22. Lancaster MA, Schroth J, Gleeson JG. Subcellular spatial regulation of canonical Wnt signalling at the primary cilium. Nat Cell Biol. 2011. doi:10.1038/ncb2259

23. Laviolette LA, Wilson J, Koller J, et al. Human Folliculin Delays Cell Cycle Progression through Late S and G2/M-Phases: Effect of Phosphorylation and Tumor Associated Mutations. PLoS One. 2013. doi:10.1371/journal.pone.0066775

24. Menko FH, van Steensel MA, Giraud S, et al. Birt-Hogg-Dubé syndrome: diagnosis and management. Lancet Oncol. 2009. doi:10.1016/S1470-2045(09)70188-3

25. Guo T, Shen Q, Ouyang R, et al. The clinical characteristics of East Asian patients with Birt-Hogg-Dubé syndrome. Ann Transl Med. 2020;8(21):1436. doi:10.21037/atm-20-1129

26. Boone PM, Scott RM, Marciniak SJ, Henske EP, Raby BA. The genetics of pneumothorax. Am J Respir Crit Care Med. 2019. doi:10.1164/rccm.201807-1212CI

27. Ray A, Paul S, Chattopadhyay E, Kundu S, Roy B. Genetic Analysis of Familial Spontaneous Pneumothorax in an Indian Family. Lung. 2015. doi:10.1007/s00408-015-9723-9

28. Köhler S, Schulz MH, Krawitz P, et al. Clinical Diagnostics in Human Genetics with Semantic Similarity Searches in Ontologies. Am J Hum Genet. 2009. doi:10.1016/j.ajhg.2009.09.003

29. Andrews S. Babraham Bioinformatics -FastQC A Quality Control tool for High Throughput Sequence Data. Soil. 1973. doi:10.1016/0038-0717(73)90093-X

30. Chang X, Wang K. Wannovar: Annotating genetic variants for personal genomes via the web. J Med Genet. 2012. doi:10.1136/jmedgenet-2012-100918

31. H L, B H, A W, et al. The Sequence Alignment/Map format and SAMtools. Bioinformatics. 2009.

32. Kim S, Scheffler K, Halpern AL, et al. Strelka2: fast and accurate calling of germline and somatic variants. Nat Methods. 2018. doi:10.1038/s41592-018-0051-x

33. Koboldt DC, Zhang Q, Larson DE, et al. VarScan 2: Somatic mutation and copy number alteration discovery in cancer by exome sequencing. Genome Res. 2012. doi:10.1101/gr.129684.111

34. Li H, Durbin R. Fast and accurate short read alignment with Burrows-Wheeler transform. Bioinformatics. 2009. doi:10.1093/bioinformatics/btp324

35. Poplin R, Ruano-Rubio V, DePristo MA, et al. Scaling accurate genetic variant discovery to tens of thousands of samples. bioRxiv. 2017. doi:10.1101/201178

36. Kircher M, Witten DM, Jain P, O’roak BJ, Cooper GM, Shendure J. A general framework for estimating the relative pathogenicity of human genetic variants. Nat Genet. 2014;46(3):310–315. doi:10.1038/ng.2892

37. Schwarz JM, Cooper DN, Schuelke M, Seelow D. Mutationtaster2: Mutation prediction for the deep-sequencing age. Nat Methods. 2014. doi:10.1038/nmeth.2890

38. Aguet F, Brown AA, Castel SE, et al. Genetic effects on gene expression across human tissues. Nature. 2017. doi:10.1038/nature24277

39. Karczewski KJ, Francioli LC, Tiao G, et al. The mutational constraint spectrum quantified from variation in 141,456 humans. bioRxiv. 2020. doi:10.1101/531210

40. Wall JD, Stawiski EW, Ratan A, et al. The GenomeAsia 100K Project enables genetic discoveries across Asia. Nature. 2019;576(7785):106–111. doi:10.1038/s41586-019-1793-z

41. Martin ER, Monks SA, Warren LL, Kaplan NL. A test for linkage and association in general pedigrees: The pedigree disequilibrium test. Am J Hum Genet. 2000;67(1):146–154. doi:10.1086/302957

42. Laskowski RA, MacArthur MW, Moss DS, Thornton JM. PROCHECK: a program to check the stereochemical quality of protein structures. J Appl Crystallogr. 1993;26(2):283–291. doi:10.1107/s0021889892009944

43. Van Zundert GCP, Rodrigues JPGLM, Trellet M, et al. The HADDOCK2.2 Web Server: User-Friendly Integrative Modeling of Biomolecular Complexes. J Mol Biol. 2016;428(4):720–725. doi:10.1016/j.jmb.2015.09.014

44. Fröhlich BA, Zeitz C, Mátyás G, et al. Novel mutations in the folliculin gene associated with spontaneous pneumothorax. Eur Respir J. 2008. doi:10.1183/09031936.00132707

45. Castel SE, Cervera A, Mohammadi P, et al. Modified penetrance of coding variants by cis-regulatory variation contributes to disease risk. doi:10.1038/s41588-018-0192-y

46. Sattler EC, Syunyaeva Z, Mansmann U, Steinlein OK. Genetic Risk Factors for Spontaneous Pneumothorax in Birt-Hogg-Dubé Syndrome. Chest. 2020;157(5):1199–1206. doi:10.1016/j.chest.2019.12.019

47. Khoo SK, Bradley M, Wong FK, Hedblad MA, Nordenskjöld M, Teh BT. Birt-Hogg-Dubé syndrome: Mapping of a novel hereditary neoplasia gene to chromosome 17p12-q11.2. Oncogene. 2001. doi:10.1038/sj.onc.1204703

48. Furuya M, Nakatani Y. Birt – Hogg – Dubé syndrome : clinicopathological features of the lung. 2013:178–186. doi:10.1136/jclinpath-2012-201200

49. Zhang X, Ma D, Zou W, et al. A rapid NGS strategy for comprehensive molecular diagnosis of Birt-Hogg-Dubé syndrome in patients with primary spontaneous pneumothorax. Respir Res. 2016. doi:10.1186/s12931-016-0377-9

50. Clausen Id L, Stein Id A, Grønbaek-Thygesen Id M, et al. Folliculin variants linked to Birt-Hogg-Dubé syndrome are targeted for proteasomal degradation. 2020. doi:10.1371/journal.pgen.1009187

