## Supplemental Methods for "Genetic Insight into Birt-Hogg-Dubé syndrome in Indian patients reveals novel mutations in *FLCN*"

### Phenotype Ontology Analysis – Phenomizer:

Detailed clinical history of each patient phenotype was correlated to Human Phenotype Ontology (HPO) terms provided in a web-based tool, Phenomizer. The tool evaluated the patient-specific HPO terms and ranked suspected Mendelian diseases from different websites (OMIM, Orphanet, and DECIPHER) by semantic similarity search. A p-value is assigned to each suspected disease based on their ranks through Benjamini-Hochberg multiple correction test.

### Targeted Amplicon NGS: DNA isolation, long PCR standardization, quantification and equimolar pooling of PCR products:

*DNA isolation and long PCR standardization*

Genomic DNA was isolated from 2ml whole blood from each participant using QIAmp DNA Isolation Midi Kit (QIAGEN Inc., Valencia, CA) as per manufacturer's protocol. DNA was amplified (Long PCR) using Roche Long Template PCR Buffer 1 (provided in the kit; Roche Holding), 10mM dNTPs, 300nM of each of forward & reverse primers, 1.5units of *Taq* polymerase for long PCR and a total of 50ng of DNA sample used for each 25µl reaction mix. Extension time for each amplicon was optimized by standard protocols. The amplicon sizes of *FLCN*, *COL3A1*, *TSC1* and *TSC2* are mentioned in Supplementary Tables S3a and S5b.

The long PCR amplifications for *FLCN* (A1 to A4), *COL3A1* (A5 and A6), *TSC1* (A7 to A11) and *TSC2* (A12 to A17) were performed by the following PCR cycle protocols -

| Amplicon | Initial Denat. | 10 cycles | | | 20 cycles | | | Final Ext. |
|---|---|---|---|---|---|---|---|---|
| | | Denat. | Ann. | Ext. | Denat. | Ann. | Ext. | |
| A1 | 94˚C for 2 mins | 94˚ for 10secs | 56˚ for 30secs | 68˚ for 5.5mins | 94˚ for 10secs | 56˚ for 30secs | 68˚ for 6mins+20s/cyc | 68˚ for 5mins |
| A2 | 94˚C for 2 mins | 94˚ for 10secs | 56˚ for 30secs | 68˚ for 5.5mins | 94˚ for 10secs | 60˚ for 30secs | 68˚ for 6mins+20s/cyc | 68˚ for 5mins |
| A3 | 94˚C for 2 mins | 94˚ for 10secs | 56˚ for 30secs | 68˚ for 4.5mins | 94˚ for 10secs | 62˚ for 30secs | 68˚ for 5mins+20s/cyc | 68˚ for 5mins |
| A4 | 94˚C for 2 mins | 94˚ for 10secs | 56˚ for 30secs | 68˚ for 5.5mins | 94˚ for 10secs | 56˚ for 30secs | 68˚ for 6mins+20s/cyc | 68˚ for 5mins |
| A5 | 94˚C for 2 mins | 94˚ for 10secs | 57˚ for 30secs | 68˚ for 6.5mins | 94˚ for 10secs | 57˚ for 30secs | 68˚ for 7mins+20s/cyc | 68˚ for 5mins |

| | | | | | | | | |
|---|---|---|---|---|---|---|---|---|
| A6 | 94˚C for 2 mins | 94˚ for 10secs | 50˚ for 30secs | 68˚ for 7.5mins | 94˚ for 10secs | 50˚ for 30secs | 68˚ for 8mins+20s/cyc | 68˚ for 5mins |
| A7 | 94˚C for 2 mins | 94˚ for 10secs | 56˚ for 30secs | 68˚ for 3.5mins | 94˚ for 10secs | 56˚ for 30secs | 68˚ for 4mins+20s/cyc | 68˚ for 5mins |
| A8 | 94˚C for 2 mins | 94˚ for 10secs | 57˚ for 30secs | 68˚ for 9mins | 94˚ for 10secs | 57˚ for 30secs | 68˚ for 9mins+20s/cyc | 68˚ for 5mins |
| A9 | 94˚C for 2 mins | 94˚ for 10secs | 57˚ for 30secs | 68˚ for 9mins | 94˚ for 10secs | 57˚ for 30secs | 68˚ for 9mins+20s/cyc | 68˚ for 5mins |
| A10 | 94˚C for 2 mins | 94˚ for 10secs | 54˚ for 30secs | 68˚ for 3.4mins | 94˚ for 10secs | 54˚ for 30secs | 68˚ for 4mins+20s/cyc | 68˚ for 5mins |
| A11 | 94˚C for 2 mins | 94˚ for 10secs | 58˚ for 30secs | 68˚ for 6mins | 94˚ for 10secs | 58˚ for 30secs | 68˚ for 6mins+20s/cyc | 68˚ for 5mins |
| A12 | 94˚C for 2 mins | 94˚ for 10secs | 53˚ for 30secs | 68˚ for 5.5mins | 94˚ for 10secs | 53˚ for 30secs | 68˚ for 6.2mins+20s/cyc | 68˚ for 5mins |
| A13 | 94˚C for 2 mins | 94˚ for 10secs | 60˚ for 30secs | 68˚ for 4mins | 94˚ for 10secs | 60˚ for 30secs | 68˚ for 4.5mins+20s/cyc | 68˚ for 5mins |
| A14 | 94˚C for 2 mins | 94˚ for 10secs | 61˚ for 30secs | 68˚ for 7.5mins | 94˚ for 10secs | 61˚ for 30secs | 68˚ for 8mins+20s/cyc | 68˚ for 5mins |
| A15 | 94˚C for 2 mins | 94˚ for 10secs | 56˚ for 30secs | 68˚ for 4.5mins | 94˚ for 10secs | 56˚ for 30secs | 68˚ for 5mins+20s/cyc | 68˚ for 5mins |
| A16 | 94˚C for 2 mins | 94˚ for 10secs | 60˚ for 30secs | 68˚ for 5mins | 94˚ for 10secs | 60˚ for 30secs | 68˚ for 5.5mins+20s/cyc | 68˚ for 5mins |
| A17 | 94˚C for 2 mins | 94˚ for 10secs | 62˚ for 30secs | 68˚ for 2.5mins | 94˚ for 10secs | 62˚ for 30secs | 68˚ for 3mins+20s/cyc | 68˚ for 5mins |

Abbreviations – Denat.: Denaturation, Ann.: Annealing, Ext.: Extension, mins: minutes, secs: seconds, s/cyc: seconds per cycle;

*Amplicon quantification and equimolar pooling*

Long PCR amplicons from 35 samples were quantified by Qubit (Invitrogen Corporation, Thermo Fisher Scientific) and nanomolar concentrations were derived using each amplicon's fragment length. Picomole concentration of each amplicon (equivalent to appx. 10µg of PCR product) were calculated and standardized for final equimolar pooling of four amplicons from 35 samples. Pooled amplicons were purified by Qiaquick PCR column purification kit as per manufacturer’s protocols (QIAGEN Inc., Valencia, CA), and later quantified in Nanodrop (Thermo Fisher Scientific). The final pooled products were diluted to 1.55ng of total DNA in 5µl of EB buffer, and quantified again by Qubit before library preparation. Library preparation was performed using the Nextera XT DNA Library Preparation kit and high throughput paired-end sequencing was done in Illumina HiSeq 2500 platform.

**Data analysis of targeted amplicons:**

Raw sequence quality was checked by FastQC followed by adaptor trimming by TrimGalore (Phred score ≥ 30). Trimmed sequence reads were mapped to the human reference genome build *(hg38)* using BWA-mem. Post alignment processing and quality filtering was done by Samtools and sequence metrics was calculated by Picard. Base quality recalibration was done by GATK4 before variant calling. Germline mutations were called by 3 variant callers - Haplotype Caller by GATK, STRELKA by Illumina, and Samtools by VarScan2. Only those variants were considered that passed through filters of strand bias and total read depth > 30. Variants were annotated using web-based wAnnovar and manually curated using Integrative Genomics Viewer Version 2 (IGV2).

***In-silico* analysis of germline variants: Damaging properties and pathogenicity prediction:**

Predictive analysis of structural and functional changes of *FLCN* due to variants was performed by SIFT and PolyPhen-2 scores by Variant Effect Predictor by Ensembl. Template CDS for *FLCN* was taken from RefSeq NM_144997, and mutant CDS were evaluated by ExPASy-Translate. Detailed analysis of the effects of any missense variants was performed by Mutation Taster-2, and splice variants by MaxENT Scan. Combined Annotated Dependent Depletion (CADD) scores were calculated to predict the pathogenicity of variants, with scores above 15 considered as pathogenic.

**Pedigree disequilibrium test (PDT) for association study:**

PDT was performed to measure linkage disequilibrium for all pedigrees. A random variable ($D$) was defined for each pedigree, given that they have informative nuclear families or informative discordant sibships. Under null hypothesis of no linkage disequilibrium, $N$ being the total number of unrelated informative pedigrees, $Di$ is taken as the summary random variable for the $i^{th}$ pedigree. With mean 0 and variance 1 under the null hypothesis, the PDT is based on the test statistic, $T$, which was calculated for all the pedigrees together to find an association between variants at *FLCN* and BHDS. Test statistic ($T$), for a one-tailed test with 5% significance, values ≥ 1.64 were considered significant.

**Homology Modelling and Protein-Protein interaction through docking using HADDOCK 2.4**

Wild-type FLCN and FNIP2 play integral roles in Rag-dependant-mTORC1 activation in lysosomes, where they directly interact with two other proteins - RRAGA and RRAGC. Due to the absence of complete crystal structures of FLCN, FNIP2, RRAGA and RRAGC; the cryo-EM structure of FLCN-FNIP2-Rag-Ragulator complex was used as a template to generate monomeric models of wild-type (FLCN, FNIP2, RRAGA, RRAGC) and FLCN mutant proteins (four exonic mutant FLCN monomers).

Interactive interfaces of wt-FLCN were obtained from the template, PDB id: *6ulg*, in which 10 residues lie in the N-terminal and 10 residues in the C-terminal.

The pdb id: *6ulg* is a hetero-nonamer, consisting of 9 chains. In this structure, FLCN interacts with 3 proteins - RRAGA, RRAGC and FNIP2. The chain L of FLCN, chain F of RRAGA, chain G of RRAGC and chain N of FNIP2 were considered for molecular docking.

In the PDB structure, 6ulg,

- FLCN (Chain L) interacts with RRAGA (Chain F) through seven amino acid residues (Arg17, Pro112, Gln116, Ser119, Arg122, His148, His231)
- FLCN (Chain L) interacts with RRAGC (Chain G) through seven different amino acid residues (Pro135, Gly136, Arg137, Gln220, Thr224, Ala225, Arg233).
- FLCN (Chain L) interacts with FNIP2 (Chain N) via 22 amino acid residues (Ile96, Ser130, Cys131, Glu132, Glu138, Pro140, Phe142, Glu146, Arg179, Lys263, Ser267, Arg268, Thr270, Glu271, Lys272, Gly276, Ala277, Val499, Glu502, Asn513, Lys516, Arg527).

HADDOCK 2.4 (High Ambiguity Driven protein-protein DOCKing) web server Guru Interface was used for macromolecular docking. It docks the interacting monomers to create docked clusters based on their biochemical and/biophysical interaction data from NMR, mutagenesis, bioinformatics predictions and cryo-electron maps. Each cluster carries a z-score denoting the similarity of the docked models. Fraction of common contacts (FCC)-based clustering was used to cluster models with a default cutoff of 0.6. Other default parameters were used for running the program and subsequent analysis. Obtained clusters were sorted according to their HADDOCK score, which is a weighted sum of van der Waals, electrostatic, desolvation and restraint violation energies together with buried surface areas. Post docking analysis was done using Pymol and interactive residues were obtained using a 5Å cut-off.

**Copy number evaluation of *FLCN*:**

*Read count normalization of NGS data by Seqmonk*

Read count normalization of *FLCN* targeted NGS data was performed by Seqmonk. Depth of coverage was calculated by GATK4. Bam files containing total read counts of *FLCN* was obtained from 20 patients and 15 asymptomatic family members. Reads within the chromosomal region of *FLCN* - chr17:17212212 to 17237618 which was divided into 5 regions, each of length 5082bp, were only considered for read length and depth normalization.

*i) FLCN*-Region 1 (chr17:17212212-17217293),

ii) *FLCN*-Region 2 (chr17:17217294-17222375),

iii) *FLCN*-Region 3 (chr17:17222376-17227457),

iv) *FLCN*-Region 4 (chr17:17227458-17232539),

v) *FLCN*-Region 5 (chr17:17232540-17237621).

Log normalized read counts in these 5 regions was calculated for each sample and used for further analysis.

*Taqman copy number assay for three exons in FLCN-*

Taqman copy number assay was performed using 3 probes for 3 exons of *FLCN* (for exons 4, 8 and 13), and one probe for *RNase P* (reference). Ct values of each sample were normalized using $\Delta Ct = Ct_{sample\ (exon\ probe)} - Ct_{sample\ (reference\ probe)}$. ΔCt values were transformed to $2^{-\Delta Ct}$ for further analysis. Parametric paired t-tests was performed for $2^{-\Delta Ct}$ values between i) Patients and Asymptomatic members, while parametric unpaired t-test was performed between ii) Patients/Asymptomatic members and Unrelated Controls for copy number analysis. Normal distribution was checked using the Kolmogorov-Smirnov tests, and all statistical analysis were carried out in SPSS.
