## Supplemental Tables for "Genetic Insight into Birt-Hogg-Dubé syndrome in Indian patients reveals novel mutations in *FLCN*"

**Supplemental Table S1: Clinically diagnosed BHDS patients and related asymptomatic members enrolled from 15 families (n=105, patients and asymptomatic members)**

| Family ID | Patient IDs with PSP or skin fibrofolliculomas | Asymptomatic member ID's | Total no. of members |
|---|---|---|---|
| F1 | F1-1, F1-2 | F1-3 to F1-8 | 8 |
| F2 | F2-9 | F2-10 to F2-12 | 4 |
| F3 | F3-13*, F3-14* | F3-15 to F3-17 | 5 |
| F4 | F4-18 | F4-19 to F4-22 | 5 |
| F5 | F5-25, F5-26, F5-28 | F5-23, F5-24, F5-27, F5-29 to F5-34 | 12 |
| F6 | F6-35* | F6-36 to F6-43 | 9 |
| F7 | F7-44*#, F7-45*, F7-46*, F7-47*, F7-48*, F7-49* | F7-50 to F7-55 | 12 |
| F8 | F8-56* | F8-57 to F8-59 | 4 |
| F9 | F9-60, F9-61, F9-62 | F9-63 to F9-66 | 7 |
| F10 | F10-67 | F10-68 to F10-69 | 3 |
| F11 | F11-70 | F11-71 to F11-76 | 7 |
| F12 | F12-77, F12-78* | F12-79 to F12-80 | 4 |
| F13 | F13-82#, F13-83, F13-84, F13-85 | F13-86 to F13-94 | 13 |
| F14 | F14-95*# | F14-96 to F14-98 | 4 |
| F15 | F15-99, F15-101 | F15-100, F15-102 to F15-106 | 8 |
| **Total** | **31** | **74** | **105** |

**Note:** All patients except F7-49 had BHDS lung phenotype (PSP or lung cysts). Patients with * also have skin fibrofolliculomas, and patients with # have renal cysts/carcinoma. Individual ID is given by Family number followed by their serial number as assigned during their enrolment in the study (eg. F1-1 is index patient from family F1). Patient IDs have been anonymised by the research group.

**Supplemental Table S2: Patient and asymptomatic members (n=35) from 11 families taken for targeted amplicon NGS of *FLCN* and *COL3A1, TSC1 & TSC2***

| Family ID | Patient ID | Asymptomatic member (AM) ID | Total |
|---|---|---|---|
| F1* | F1-1* | - | 1 |
| F2 | F2-9 | F2-10, F2-12 | 3 |
| F3 | F3-13, F3-14 | F3-15 | 3 |
| F4 | F4-18 | F4-19 | 2 |
| F5 | F5-25, F5-26, F5-28 | F5-23, F5-24 | 5 |
| F6 | F6-35 | F6-36 to F6-40 | 6 |
| F7 | F7-44 to F7-48 | 0 | 5 |
| F8 | F8-56 | F8-57, F8-58 | 3 |
| F9 | F9-60, F9-61, F9-62 | F9-63 | 4 |
| F10 | F10-67 | F10-68 | 2 |
| F11** | F11-70** | - | 1 |
| **Total** | **20** | **15** | **35** |

*Result of *FLCN* mutation from patient F1-1 of family F1 has been published in an earlier study (Ray *et al.*, *Lung, 2015*), therefore mutation data from F1-1 taken as positive control. Other family members from family F1 were not included in this sequencing method.

** Patient F11-70 is the only individual included from family F11 due to logistic constraints

**Supplemental Table S3a: Genomic coordinates and primer sequences for *FLCN* long PCR encompassing exons and UTRs, used in targeted amplicon NGS**

| Amplicon name | Genomic Coordinates | Sequences (5' to 3') | Amplicon Length (bp) | UTR's and exons included |
|---|---|---|---|---|
| FLN – A1 | chr17:17,237,734 to 17,231,271 | *CGAGTTCTGCAACCAACCTC* | 6464 | 3'UTR and exons 1, 2, 3 |
| | | *GTCCTGAGTGTTTGCTAGGC* | | |
| FLN – A2 | chr17:17,232,245 to 17,225,868 | *GCAAGGGGTACTCTGAGCAG* | 6378 | Exons 3, 4, 5 |
| | | *GAGACCCTGACACAAAAGAAGG* | | |
| FLN – A3 | chr17:17,223,045 to 17,217,787 | *CCTTGTTTGTCTCAGCTCATTG* | 5259 | Exons 7, 8, 9 |
| | | *GACACCTTAGGAGAGTCCAGC* | | |
| FLN – A4 | chr17:17,217,807 to 17,211,688 | *GCTGGACTCTCCTAAGGTGTC* | 6120 | Exons 10, 11, 12, 13, 14, and 5'UTR |
| | | *GCATAGGTACTCAACAGATGTCC* | | |

**Supplemental Table S3b: Primers to amplify 14 exons and their flanking intronic regions (50bp) of *FLCN* by Sanger Sequencing method**

| PRIMERS | SEQUENCES |
|---|---|
| Exon 1 (UTR) | *5'- GTGTTGGGTGGTGGTACG-3'* |
| | *5'- CCAACGAAAACTCGGACA -3'* |
| Exon 2 (UTR) | *5'- CTAAGCCATTCAGAACCCTG -3'* |
| | *5'- GGCATTAAACTGCGAAAAGG -3'* |
| Exon 3 (UTR) | *5'- TCCCTCTCTGACTCCCACAC -3'* |
| | *5'- GCCAAAGCCGCTAACTCTAG -3'* |
| Exon 4 | *5'- TCTCCTGGGCAGGAAGTC -3'* |
| | *5'- GTCAGGATGAGCGGAAAGAA -3'* |
| Exon 5 | *5'- GCCCTGCTTCCCAACTAA -3'* |
| | *5'- GCCCTGAGAGAGGACCAGT -3'* |
| Exon 6 | *5'- TTGTGCCAGCTGACTCTG -3'* |
| | *5'- CCAGCTCTGAAGCCAAGA -3'* |
| Exon 7 | *5'- GTGGGACTGATCCTCCAG -3'* |
| | *5'- CAGAGGCAGCAAGCAAAC -3'* |
| Exon 8 | *5'- GTGAGCGTCAGGTTTGCTTT -3'* |
| | *5'- CTGCCAGGAGAGCAGACAG -3'* |
| Exon 9 | *5'- GGGCTGAAGTCACAGGAT -3'* |
| | *5'- CCATGGGATGCCAACTAT -3'* |
| Exon 10 | *5'- GTCACGCTGAAAGCACTG -3'* |
| | *5'- ACATCATCAGACCAGACCC -3'* |
| Exon 11 | *5'- CACTGTGGGCTGAGAGTCTG -3'* |
| | *5'- CCTCTCCACAACCCATGA -3'* |
| Exon 12-13 | *5'- CCACTGACCTGGGATGAG -3'* |
| | *5'- GGCCCAGCTCCTCTTTTG -3'* |
| Exon 14 | *5'- GCTCGAGGGATTGTGCTG -3'* |
| | *5'- GCCCAGAAACTCTTGCTG -3'* |
| Exon 14 (UTR) | *5'- TCCTTGGAGAGACGACTAGG -3'* |
| | *5'- CCGGCTGACTCACTGGTAT -3'* |

**Supplemental Table S4: Patients and asymptomatic members in 4 families taken for Taqman *FLCN* copy number assays**

| **Family ID** | **Patient ID** | **Asymptomatic member (AM) ID** | **Total individuals** |
|---|---|---|---|
| F3 | F3-13, F3-14 | F3-15 to F3-17 | 5 |
| F4 | F4-18 | F4-19 to F4-22 | 5 |
| F9 | F9-60, F9-61, F9-62 | F9-63 to 66 | 7 |
| F10 | F10-67 | F10-68 & F10-69 | 3 |
| **4** | **7** | **13** | **20** |

**Supplemental Table S5a: Patients and asymptomatic members in 5 families taken for detection of germline mutations in three genes - *SERPINA1*, *MTHFR* and *CBS* by Sanger sequencing**

| **Family ID** | **Patient ID** | **Asymptomatic member (AM) ID** | **Total individuals** |
|---|---|---|---|
| F6 | F6-35 | F6-36 to F6-40 | 6 |
| F7 | F7-44 to F7-48 | F7-54, F7-55 | 7 |
| F8 | F8-56 | F8-57, F8-58 | 3 |
| F9 | F9-60, F9-61, F9-62 | F9-63 to F9-66 | 7 |
| F10 | F10-67 | F10-68 | 2 |
| **5** | **11** | **14** | **25** |

**Supplemental Table S5b: PCR primer details for amplification *COL3A1*, *TSC1* and *TSC2* in targeted amplicon NGS study**

| Amplicon name | Genomic Coordinates of PCR Primers | Amplicon Size | Exons included |
|---|---|---|---|
| C31 – A5 | chr2: 188985986-188993765 | 7780 | exons 5 to 15 |
| C31 – A6 | chr2: 188992492-189000916 | 8425 | exons 15 to 32 |
| TC1 – A7 | chr9: 132933666-132938298 | 4633 | exon 2 |
| TC1 – A8 | chr9: 132925801-132935485 | 9685 | exons 2 to 4 |
| TC1 – A9 | chr9: 132911721-132922255 | 10535 | exons 7 to 9 |
| TC1 – A10 | chr9: 132896920-132902086 | 5167 | exons 19 to 22 |
| TC1 – A11 | chr9: 132890784-132898073 | 7290 | exons 21 to 23 |
| TS1 – A12 | chr16: 2047032-2053669 | 6638 | exons 1 to 4 |
| TS1 – A13 | chr16: 2052925-2058416 | 5492 | exons 4 to 9 |
| TS1 – A14 | chr16: 2057674-2066119 | 8446 | exons 10 to 16 |
| TS1 – A15 | chr16: 2077141-2082672 | 5532 | exons 26 to 32 |
| TS1 – A16 | chr16: 2081133-2087213 | 6081 | exons 31 to 38 |
| TS1 – A17 | chr16: 2085606-2089401 | 3796 | exons 37 to 42 |

**Note:** Abbreviations: C31: Primers for *COL3A1*, TC1: Primers for *TSC1*, TS1: Primers for *TSC2*. About 15kb of 38kb of *COL3A1* (25 of 51 exons and flanking introns), 23.5kb of 53kb of *TSC1* (12 of 42 exons and flanking introns), and 31.3kb of 41kb of *TSC2* (33 of 42 exons and flanking introns) were amplified and sequenced. PCR protocols are mentioned in Supplemental Methods.

**Supplemental Table S6: Thirty one patients from 15 families with major or minor diagnostic clinical criteria for BHDS**

| S. No | Family ID | Sample ID | Sex | Age Range (yr) | *Major Criteria* | *Minor Criteria* | | |
|---|---|---|---|---|---|---|---|---|
| | | | | | **Fibrofolli-culomas** | **Pneumo-thorax or lung cysts** | **Renal cell carcinoma/ renal cysts** | **First degree relative** |
| 1 | F1 | F1- 1 | F | 31-35 | absent | present | absent | present |
| 2 | | F1- 2 | F | 31-35 | absent | present | absent | present |
| 3 | F2 | F2-9* | M | 46-50 | absent | present | absent | absent |
| 4 | F3 | F3-13 | M | 41-45 | **present** | present | absent | present |
| 5 | | F3-14 | M | 41-45 | **present** | present | absent | present |
| 6 | F4 | F4-18* | M | 41-45 | absent | present | absent | absent |
| 7 | F5 | F5-25 | F | 36-40 | absent | present | absent | present |
| 8 | | F5-26 | M | 36-40 | absent | present | absent | present |
| 9 | | F5-28 | M | 31-35 | absent | present | absent | present |
| 10 | F6 | F6-35 | M | 61-65 | **present** | present | absent | absent |
| 11 | F7 | F7-44 | M | 56-60 | **present** | present | **present** | present |
| 12 | | F7-45 | F | 61-65 | **present** | present | absent | present |
| 13 | | F7-46 | M | 61-65 | absent | present | absent | present |
| 14 | | F7-47 | F | 51-55 | **present** | present | absent | present |
| 15 | | F7-48 | F | 75-80 | **present** | present | absent | present |
| 16 | | F7-49 | M | 85-90 | **present** | unknown | absent | present |
| 17 | F8 | F8-56 | F | 15-20 | **present** | present | absent | absent |
| 18 | F9 | F9-60 | M | 36-40 | absent | present | absent | present |
| 19 | | F9-61 | F | 61-65 | absent | present | absent | present |
| 20 | | F9-62 | M | 36-40 | absent | present | absent | present |
| 21 | F10 | F10-67* | F | 56-60 | absent | present | absent | absent |
| 22 | F11 | F11-70* | F | 41-45 | absent | present | absent | absent |
| 23 | F12 | F12-77 | F | 71-75 | **present** | present | absent | present |
| 24 | | F12-78 | M | 41-45 | absent | present | absent | present |
| 25 | F13 | F13-82 | M | 56-60 | absent | present | **present** | present |
| 26 | | F13-83 | M | 51-55 | absent | present | absent | present |
| 27 | | F13-84 | M | 46-50 | absent | present | absent | present |
| 28 | | F13-85 | M | 36-40 | absent | present | absent | present |
| 29 | F14 | F14-95 | F | 31-35 | **present** | present | **present** | Absent |
| 30 | F15 | F15-99 | M | 61-65 | absent | present | absent | present |
| 31 | | F15-101 | F | 31-35 | absent | present | absent | present |

* Patients fulfilling only one criterion (lung phenotype); Patient F11-70 has second degree relatives with BHDS. In our cohort, 4 patients only have one criterion (lung phenotype), but had BHDS-specific lung cysts or repeated pneumothoraces. BHDS diagnostic criteria laid out by *Menko et al., 2009* stated, for a patient to be considered as BHDS, they should qualify for either one major criterion (skin fibrofolliculomas or *FLCN* pathogenic mutation) or ≥ 2 minor criteria, or 1 major & 1 minor criterion.

M: Male, F: Female. Bold indicates presence of skin fibrofolliculomas, and renal cysts/carcinoma.

**Supplemental Table S7: Phenotype Ontology Analysis using Phenomizer for 31 patients**

| Family ID | Patient ID | p-value | Score | OMIM | Description | Gene |
|---|---|---|---|---|---|---|
| F1 | F1-1 | **0.0053** | 4.4332 | 173600 | Primary Spontaneous Pneumothorax | *FLCN* |
| | F1-2 | **0.0053** | 3.7429 | 173600 | Primary Spontaneous Pneumothorax | *FLCN* |
| F2 | F2-9 | **0.0053** | 3.7429 | 173600 | Primary Spontaneous Pneumothorax | *FLCN* |
| F3 | F3-13 | **0.0027** | 5.3443 | 135150 | Birt-Hogg-Dubé syndrome | *FLCN* |
| | F3-14 | 0.3057 | 4.2794 | 135150 | Birt-Hogg-Dubé syndrome | *FLCN* |
| F4 | F4-18 | **0.016** | 3.4511 | 173600 | Primary Spontaneous Pneumothorax | *FLCN* |
| F5 | F5-25 | **0.016** | 3.4511 | 173601 | Primary Spontaneous Pneumothorax | *FLCN* |
| | F5-26 | 0.1646 | 4.7636 | 158310 | Hereditary Mucoepthelial Dysplasia | - |
| | F5-28 | 0.1646 | 4.7636 | 158310 | Hereditary Mucoepthelial Dysplasia | - |
| F6 | F6-35 | **0.016** | 2.8072 | 173600 | Primary Spontaneous Pneumothorax | *FLCN* |
| F7 | F7-44 | **0.0027** | 5.5242 | 135150 | Birt-Hogg-Dubé syndrome | *FLCN* |
| | F7-45 | **0.0027** | 5.3443 | 135150 | Birt-Hogg-Dubé syndrome | *FLCN* |
| | F7-46 | **0.0053** | 3.7429 | 173600 | Primary Spontaneous Pneumothorax | *FLCN* |
| | F7-47 | **0.0053** | 5.0544 | 135150 | Birt-Hogg-Dubé syndrome | *FLCN* |
| | F7-48 | **0.0027** | 5.3443 | 135150 | Birt-Hogg-Dubé syndrome | *FLCN* |
| | F7-49 | 0.2532 | 5.4567 | 135150 | Birt-Hogg-Dubé syndrome | *FLCN* |
| F8 | F8-56 | **0.0053** | 6.0976 | 135150 | Birt-Hogg-Dubé syndrome | *FLCN* |
| F9 | F9-60 | 0.1646 | 4.7636 | 158310 | Hereditary Mucoepthelial Dysplasia | - |
| | F9-61 | 0.1646 | 4.7636 | 158310 | Hereditary Mucoepthelial Dysplasia | - |
| | F9-62 | **0.0266** | 4.4518 | 610913 | ILD due to surfactant protein C deficiency | SFTPC |
| F10 | F10-67 | **0.016** | 2.8072 | 173600 | Primary Spontaneous Pneumothorax | *FLCN* |
| F11 | F11-70 | **0.0053** | 4.4332 | 173600 | Primary Spontaneous Pneumothorax | *FLCN* |
| F12 | F12-77 | **0.0053** | 4.652 | 173600 | Primary Spontaneous Pneumothorax | *FLCN* |
| | F12-78 | **0.0053** | 3.7429 | 173600 | Primary Spontaneous Pneumothorax | *FLCN* |
| F13 | F13-82 | **0.0018** | 4.3871 | 135150 | Birt-Hogg-Dubé syndrome | *FLCN* |
| | F13-83 | **0.0053** | 4.652 | 173600 | Primary Spontaneous Pneumothorax | *FLCN* |
| | F13-84 | **0.0053** | 4.652 | 173600 | Primary Spontaneous Pneumothorax | *FLCN* |
| | F13-85 | **0.0053** | 4.4332 | 173600 | Primary Spontaneous Pneumothorax | *FLCN* |
| F14 | F14-95 | **0.0053** | 5.5034 | 135150 | Birt-Hogg-Dubé syndrome | *FLCN* |
| F15 | F15-99 | 0.1646 | 4.7636 | 158310 | Hereditary Mucoepthelial Dysplasia | - |
| | F15-101 | **0.0053** | 3.5465 | 173600 | Primary Spontaneous Pneumothorax | *FLCN* |

Note: bold p-values means it is less than 0.05, ILD: interstitial lung disease

**Supplemental Table S8: Total Read Count for 20 patients and 15 asymptomatic members obtained from *FLCN* targeted amplicon NGS data (post quality filters)**

| Sample ID | Patient/Asymptomatic member | Total Read Count (SeqMonk) |
|---|---|---|
| F1-1 | Patient | 7492195 |
| F2-9 | Patient | 7145316 |
| F2-10 | Asymptomatic member | 6742544 |
| F2-12 | Asymptomatic member | 5171128 |
| F3-13 | Patient | 5928218 |
| F3-14 | Patient | 5510277 |
| F3-15 | Asymptomatic member | 6196936 |
| F4-18 | Patient | 7614903 |
| F4-19 | Asymptomatic member | 8028362 |
| F5-23 | Asymptomatic member | 5132578 |
| F5-24 | Asymptomatic member | 7808531 |
| F5-25 | Patient | 8367280 |
| F5-26 | Patient | 7863282 |
| F5-28 | Patient | 8663063 |
| F6-35 | Patient | 7957945 |
| F6-36 | Asymptomatic member | 6794231 |
| F6-37 | Asymptomatic member | 6825705 |
| F6-38 | Asymptomatic member | 7624874 |
| F6-39 | Asymptomatic member | 5776490 |
| F6-40 | Asymptomatic member | 6612516 |
| F7-44 | Patient | 9033619 |
| F7-45 | Patient | 7346030 |
| F7-46 | Patient | 6655348 |
| F7-47 | Patient | 7955265 |
| F7-49 | Patient | 7379168 |
| F8-56 | Patient | 6311258 |
| F8-57 | Asymptomatic member | 9789782 |
| F8-58 | Asymptomatic member | 8156624 |
| F9-60 | Patient | 6556172 |
| F9-61 | Patient | 6174414 |
| F9-62 | Patient | 8032191 |
| F9-63 | Asymptomatic member | 6827487 |
| F10-67 | Patient | 6919623 |
| F10-68 | Asymptomatic member | 8557839 |
| F11-70 | Patient | 7266443 |

**Note:** The reads were aligned to human reference (hg38) and after various quality filters, an average of 7.2 million reads was obtained, while the average depth of coverage of all samples was ~18,000x.

**Supplemental Table S9a: Non-pathogenic/non-coding SNV's in *FLCN* found by NGS**

(Presence of mutations in **brown in** 3' UTR, **green in** intron and **blue in** 5'UTR)

| S. No. | COORD | ID | HGVS | Annotation | REF | ALT | Benign/ published reports | F2 | F3 | F4 | F5 | F6 | F7 | F8 | F9 | F10 | F1-1 | F11-70 |
|---|---|---|---|---|---|---|---|---|---|---|---|---|---|---|---|---|---|---|
| 1 | 17212251 | *rs1451192210* | *c.*1404C>T* | 3' UTR | *G* | *A* | | | | | | | | | | | | |
| 2 | 17212252 | *rs7218795* | *c.*1403T>C* | 3' UTR | *A* | *G* | benign | | | | | | | | | | | |
| 3 | 17212319 | *rs7218992* | *c.*1336G>A* | 3' UTR | *C* | *A* | benign | | | | | | | | | | | |
| 4 | 17213098 | *rs3803761* | *c.*557T>C* | 3' UTR | *A* | *G* | benign | | | | | | | | | | | |
| 5 | 17213230 | *rs7224335* | *c.*425G>A* | 3' UTR | *C* | *T* | benign | | | | | | | | | | | |
| 6 | 17213262 | *rs12602675* | *c.*393G>A* | 3' UTR | *C* | *T* | benign | | | | | | | | | | | |
| 7 | 17213299 | *rs7224474* | *c.*356G>T* | 3' UTR | *C* | *A* | benign | | | | | | | | | | | |
| 8 | 17214195 | *rs8068606* | *c.1539-339A>C* | intron variant | *T* | *G* | | | | | | | | | | | | |
| 9 | 17214314 | *rs8067893* | *c.1539-458C>T* | intron variant | *G* | *A* | | | | | | | | | | | | |
| 10 | 17214401 | - | *c.1539-545G>A* | intron variant | *C* | *T* | | | | | | | | | | | | |
| 11 | 17215128 | *rs34235236* | *c.1433-38A>G* | intron variant | *T* | *C* | benign | | | | | | | | | | | |
| 12 | 17215375 | *rs34311146* | *c.1301-59C>T* | intron variant | *G* | *A* | published | | | | | | | | | | | |
| 13 | 17215386 | *rs544930629* | *c.1301-70C>T* | intron variant | *G* | *A* | | | | | | | | | | | | |
| 14 | 17215879 | *rs572265105* | *c.1300+501A>G* | intron variant | *T* | *C* | | | | | | | | | | | | |
| 15 | 17216890 | *rs7208065* | *c.1176+179A>G* | intron variant | *T* | *C* | published | | | | | | | | | | | |
| 16 | 17217030 | *rs41424546* | *c.1176+39G>A* | intron variant | *C* | *T* | published | | | | | | | | | | | |
| 17 | 17217354 | *rs4985705* | *c.1063-172C>G* | intron variant | *G* | *C* | published | | | | | | | | | | | |
| 18 | 17217498 | *rs4985752* | *c.1063-316A>G* | intron variant | *T* | *C* | | | | | | | | | | | | |
| 19 | 17218310 | *rs8080386* | *c.1062+709A>G* | intron variant | *T* | *C* | | | | | | | | | | | | |
| 20 | 17218972 | *rs8065572* | *c.1062+47G>A* | intron variant | *C* | *T* | | | | | | | | | | | | |
| 21 | 17219013 | *rs8065832* | *c.1062+6C>T* | intron variant | *G* | *A* | benign | | | | | | | | | | | |
| 22 | 17219819 | *rs2018781* | *c.872-610C>G* | intron variant | *G* | *C* | published | | | | | | | | | | | |
| 23 | 17220853 | *rs41323249* | *c.871+684G>A* | intron variant | *C* | *T* | published | | | | | | | | | | | |
| 24 | 17221311 | *rs41400246* | *c.871+226G>A* | intron variant | *C* | *T* | published | | | | | | | | | | | |

| | | | | | | | |
|---|---|---|---|---|---|---|---|
| 25 | 17221501 | *rs3744124* | *c.871+36G>A* | intron variant | *C* | *T* | benign |
| 26 | 17222119 | *rs1708622* | *c.779+382T>C* | intron variant | *A* | *G* | |
| 27 | 17222281 | *rs1736221* | *c.779+220T>C* | intron variant | *A* | *G* | |
| 28 | 17222727 | *rs2292527* | *c.619-66C>T* | intron variant | *G* | *A* | |
| 29 | 17226117 | *rs41525346* | *c.396+59T>C* | intron variant | *A* | *G* | benign |
| 30 | 17226610 | *rs1708620* | *c.250-288G>A* | intron variant | *C* | *T* | published |
| 31 | 17226729 | *rs1708619* | *c.250-407G>A* | intron variant | *C* | *T* | |
| 32 | 17226923 | *rs1736216* | *c.250-601C>T* | intron variant | *G* | *A* | |
| 33 | 17226931 | *rs6502565* | *c.250-609G>A* | intron variant | *C* | *T* | |
| 34 | 17226956 | *rs76319098* | *c.250-634G>A* | intron variant | *C* | *T* | |
| 35 | 17227493 | *rs373903620* | *c.249+396G>A* | intron variant | *C* | *T* | |
| 36 | 17227704 | *rs55836267* | *c.249+185A>C* | intron variant | *T* | *G* | |
| 37 | 17228404 | *rs75336342* | *c.-24-243G>A* | intron variant | *C* | *T* | |
| 38 | 17228555 | *rs1708618* | *c.-24-394A>G* | intron variant | *T* | *C* | |
| 39 | 17228589 | *rs538804082* | *c.-24-428G>T* | intron variant | *C* | *A* | |
| 40 | 17228852 | *rs79717038* | *c.-24-691C>T* | intron variant | *G* | *A* | |
| 41 | 17228888 | *rs1736215* | *c.-24-727C>T* | intron variant | *G* | *A* | published |
| 42 | 17229019 | *rs1736214* | *c.-24-858G>C* | intron variant | *C* | *G* | |
| 43 | 17229099 | *rs565915341* | *c.-24-938C>T* | intron variant | *G* | *A* | |
| 44 | 17229304 | *rs1708617* | *c.-24-1143T>C* | intron variant | *A* | *G* | |
| 45 | 17229807 | *rs1613416* | *c.-24-1646C>T* | intron variant | *G* | *A* | |
| 46 | 17230965 | *rs12602831* | *c.-25+829C>T* | intron variant | *G* | *T* | |
| 47 | 17231193 | *rs12602871* | *c.-25+601C>T* | intron variant | *G* | *A* | |
| 48 | 17231214 | *rs1736213* | *c.-25+580A>C* | intron variant | *T* | *G* | published |
| 49 | 17231694 | *rs1736212* | *c.-25+100C>G* | intron variant | *G* | *C* | published |
| 50 | 17232108 | *rs1736211* | *c.-113-226T>C* | intron variant | *A* | *G* | |
| 51 | 17233101 | *rs76045368* | *c.-227-200G>A* | intron variant | *C* | *T* | |
| 52 | 17233301 | *rs2349865* | *c.-227-400T>A* | intron variant | *A* | *T* | |
| 53 | 17233347 | *rs34518797* | *c.-227-446G>A* | intron variant | *C* | *T* | |
| 54 | 17233379 | *rs11078378* | *c.-227-478C>G* | intron variant | *G* | *C* | |

| | | | | | | | |
|---|---|---|---|---|---|---|---|
| 55 | 17233577 | *rs10459910* | *c.-227-676G>A* | intron variant | *C* | *T* | |
| 56 | 17233992 | *rs8079562* | *c.-227-1091C>G* | intron variant | *G* | *A* | |
| 57 | 17234361 | *rs375429199* | *c.-227-1460C>T* | intron variant | *G* | *A* | |
| 58 | 17234497 | *rs8079971* | *c.-227-1596T>C* | intron variant | *A* | *G* | |
| 59 | 17234845 | *rs8065774* | *c.-227-1944G>A* | intron variant | *C* | *T* | |
| 60 | 17235067 | *rs8066090* | *c.-228+1845T>C* | intron variant | *A* | *G* | |
| 61 | 17235918 | *rs41337846* | *c.-228+994A>G* | intron variant | *T* | *C* | published |
| 62 | 17236983 | *rs1708629* | *c.-299C>T* | 5' UTR | *G* | *A* | benign |
| 63 | 17236986 | *rs41345949* | *c.-302G>A* | 5' UTR | *C* | *T* | benign |
| 64 | 17237171 | *rs1736209* | *c.-287G>C* | 5' UTR | *C* | *G* | benign |

**Note:** COORD: chromosomal position. Coloured boxes indicate presence of the variant in that family. 3’ UTR variants are denoted by brown, 5’ UTR variants in blue, and intronic variants in green.

**Supplemental Table S9b: Non-coding indels in *FLCN* found by NGS** (Presence of mutation in **green:** intron)

<table>
<tr><th>S. No.</th><th>COORD</th><th>ID</th><th>HGVS</th><th>Annotation</th><th>REF</th><th>ALT</th><th>Benign/ published reports</th><th>F2</th><th>F3</th><th>F4</th><th>F5</th><th>F6</th><th>F7</th><th>F8</th><th>F9</th><th>F 10</th><th>F1-1</th><th>F 11-70</th></tr>
<tr><td>1</td><td>17214142</td><td>rs10681411</td><td>c.1539-290_ 1539-287dup</td><td>intronic</td><td>G</td><td>-/G</td><td>-</td><td></td><td style="background:green"></td><td style="background:green"></td><td></td><td></td><td></td><td style="background:green"></td><td></td><td></td><td></td><td style="background:green"></td></tr>
<tr><td>2</td><td>17214701</td><td>rs146159727</td><td>c.1538+281_ 1538+284del</td><td>intronic</td><td>CATT</td><td>CATT/-</td><td>-</td><td></td><td style="background:green"></td><td></td><td></td><td></td><td></td><td></td><td></td><td></td><td></td><td></td></tr>
<tr><td>3</td><td>17227281</td><td>rs113749420</td><td>c.249+604_ 249+607del</td><td>intronic</td><td>ACCC</td><td>ACCC/-</td><td>-</td><td style="background:green"></td><td></td><td style="background:green"></td><td style="background:green"></td><td></td><td></td><td style="background:green"></td><td></td><td></td><td></td><td></td></tr>
<tr><td>4</td><td>17228584</td><td>rs200365194</td><td>c.-24-425_ -24-424del</td><td>intronic</td><td>GG</td><td>GG/-</td><td>-</td><td></td><td></td><td style="background:green"></td><td></td><td style="background:green"></td><td></td><td></td><td></td><td></td><td></td><td style="background:green"></td></tr>
<tr><td>5</td><td>17228592</td><td>rs879808064</td><td>c.-24-435_ -24-432del</td><td>intronic</td><td>GG</td><td>GG/-</td><td>-</td><td></td><td></td><td style="background:green"></td><td></td><td style="background:green"></td><td></td><td></td><td></td><td></td><td></td><td></td></tr>
<tr><td>6</td><td>17229504</td><td>rs66460366</td><td>c.-24-1345_ -24-1344del</td><td>intronic</td><td>AT</td><td>AT/-</td><td>-</td><td style="background:green"></td><td style="background:green"></td><td style="background:green"></td><td></td><td style="background:green"></td><td style="background:green"></td><td style="background:green"></td><td style="background:green"></td><td style="background:green"></td><td style="background:green"></td><td style="background:green"></td></tr>
</table>

**Note:** COORD: chromosomal position. Coloured boxes indicate presence of the variant in that family. 3’ UTR variants are denoted by brown, 5’ UTR variants in blue, and intronic variants in green

**Supplemental Table S10a: Normalized gene expression values (median) of *FLCN* associated with 45 different SNPs (found in our cohort) common with *FLCN* SNPs in e-QTL data of Gtex database in Lung and Skin (exposed and unexposed) tissues**

| S. No. | COORD | ID | HGVS | REF | ALT - FREQ (dbSNP) | LUNG genotypes (normalized expression median values) | | | |
|---|---|---|---|---|---|---|---|---|---|
| | | | | | | p-value | REF | HET | ALT |
| 1 | 17236983 | *rs1708629* | 5' UTR variant | G | A=0.41 | 1.90E-82 | -0.65(144) | -0.014(241) | 0.69(130) |
| 2 | 17236986 | *rs41345949* | 5' UTR variant | C | T=0.05 | 1.10E-07 | -0.06(440) | 0.27(69) | N.A |
| 3 | 17237171 | *rs1736209* | 5' UTR variant | C | G=0.72 | 7.30E-23 | -0.65(33) | -0.20(220) | 0.28(262) |
| 4 | 17212252 | *rs7218795* | 3' UTR variant | A | G=0.66 | 8.20E-25 | -0.67(40) | -0.22(213) | 0.34 (262) |
| 5 | 17212319 | *rs7218992* | 3' UTR variant | C | A=0.06 | 8.90E-14 | 0.126(384) | -0.37 (119) | -0.91 (12) |
| 6 | 17213098 | *rs3803761* | 3' UTR variant | A | G=0.65 | 3.00E-25 | -0.67(42) | -0.21(213) | 0.32 (260) |
| 7 | 17213262 | *rs12602675* | 3' UTR variant | C | T=0.08 | 0.00011 | 0.014(478) | -0.29(37) | N.A |
| 8 | 17216890 | *rs7208065* | intron variant | T | C=0.48 | 7.10E-86 | -0.782(120) | -0.02(260) | 0.724(135) |
| 9 | 17217030 | *rs41424546* | intron variant | C | T=0.108 | 6.90E-14 | 0.13(396) | -0.37(109) | -0.4(10) |
| 10 | 17217354 | *rs4985705* | intron variant | G | C=0.47 | 4.50E-88 | -0.79(121) | -0.014(257) | 0.71(137) |
| 11 | 17217498 | *rs4985752* | intron variant | T | C= 0.41 | 2.80E-81 | -0.66(144) | 0 (243) | 0.71(128) |
| 12 | 17218310 | *rs8080386* | intron variant | T | C=0.60 | 2.10E-55 | -0.79(83) | -0.08(266) | 0.5(166) |
| 13 | 17219013 | *rs8065832* | intron variant | G | A=0.43 | 9.30E-65 | -0.66(147) | 0.08 (259) | 0.65 (109) |
| 14 | 17219819 | *rs2018781* | intron variant | G | C=0.73 | 4.90E-21 | -0.65(25) | -0.26(211) | 0.27(279) |
| 15 | 17221501 | *rs3744124* | intron variant | C | T=0.12 | 2.40E-09 | 0.07 (452) | -0.44 (62) | N.A |
| 16 | 17222119 | *rs1708622* | intron variant | A | G=0.74 | 8.10E-20 | -0.65(22) | -0.26(209) | 0.25(284) |
| 17 | 17222281 | *rs1736221* | intron variant | A | G=0.8 | 2.10E-55 | -0.79(83) | -0.089(266) | 0.57(166) |
| 18 | 17226610 | *rs1708620* | intron variant | C | T=0.43 | 3.60E-66 | -0.65(146) | 0.08(258) | 0.65(111) |
| 19 | 17226729 | *rs1708619* | intron variant | C | T= 0.44 | 9.90E-66 | -0.65(145) | 0.08(259) | 0.65(111) |
| 20 | 17226923 | *rs1736216* | intron variant | G | A=0.70 | 6.50E-11 | -0.46(40) | -0.12(224) | 0.16(251) |
| 21 | 17226931 | *rs6502565* | intron variant | C | T=0.13 | 1.80E-09 | 0.068(451) | -0.44(62) | N.A |
| 22 | 17226956 | *rs76319098* | intron variant | C | T=0.13 | 1.80E-09 | 0.05(466) | -0.47(47) | N.A |
| 23 | 17228555 | *rs1708618* | intron variant | T | C=0.47 | 1.10E-96 | -0.78(126) | 0.009(253) | 0.65(136) |
| 24 | 17228852 | *rs79717038* | intron variant | G | A=0.13 | 2.70E-08 | 0.06 (464) | -0.47(49) | N.A |

| | | | | | | | | | |
|---|---|---|---|---|---|---|---|---|---|
| 25 | 17228888 | *rs1736215* | intron variant | G | A=0.46 | 2.30E-90 | -0.76(119) | -0.02(256) | 0.69(140) |
| 26 | 17229019 | *rs1736214* | intron variant | C | G=0.46 | 2.10E-88 | -0.8(118) | -0.014(257) | 0.65(140) |
| 27 | 17229304 | *rs1708617* | intron variant | A | G=0.72 | 9.10E-23 | -0.65(34) | -0.20(218) | 0.29(263) |
| 28 | 17229807 | *rs1613416* | intron variant | G | A=0.47 | 2.30E-90 | -0.78(120) | -0.01(254) | 0.66(141) |
| 29 | 17231214 | *rs1736213* | intron variant | T | G=0.48 | 2.00E-90 | -0.79(119) | -0.01(256) | 0.69(140) |
| 30 | 17231694 | *rs1736212* | intron variant | G | C=0.72 | 9.10E-23 | -0.65(34) | -0.20(218) | 0.29(263) |
| 31 | 17232108 | *rs1736211* | intron variant | A | G=0.74 | 1.70E-20 | -0.65(24) | -0.24(203) | 0.23(288) |
| 32 | 17233301 | *rs2349865* | intron variant | A | T=0.25 | 8.80E-23 | -0.65(34) | -0.20(219) | 0.28 (262) |
| 33 | 17233379 | *rs11078378* | intron variant | G | C=0.74 | 3.90E-20 | -0.65(23) | -0.24(205) | 0.22(287) |
| 34 | 17214314 | *rs8067893* | intron variant | G | A=0.11 | 2.80E-12 | 0.134(340) | -0.255(151) | -0.52 (24) |
| 35 | 17220853 | *rs41323249* | intron variant | C | T=0.12 | 2.60E-14 | 0.14(359) | -0.34(139) | -0.50(17) |
| 36 | 17221311 | *rs41400246* | intron variant | C | T=0.10 | 2.10E-14 | 0.13(393) | -0.37(112) | -0.4 (10) |
| 37 | 17226117 | *rs41525346* | intron variant | A | G=0.08 | 9.90E-08 | 0.08(436) | -0.43(76) | N.A |
| 38 | 17227704 | *rs55836267* | intron variant | T | G= 0.12 | 2.60E-14 | 0.14(359) | -0.34(139) | -0.50(17) |
| 39 | 17230965 | *rs12602831* | intron variant | G | T=0.25 | 7.10E-24 | 0.20(356) | -0.34(140) | -0.42(19) |
| 40 | 17231193 | *rs12602871* | intron variant | G | A=0.25 | 7.10E-24 | 0.20(356) | -0.34(140) | -0.42(19) |
| 41 | 17233101 | *rs76045368* | intron variant | C | T=0.25 | 3.30E-25 | 0.24(331) | -0.35(160) | -0.46(24) |
| 42 | 17233347 | *rs34518797* | intron variant | C | T=0.25 | 3.30E-25 | 0.24(331) | -0.35(160) | -0.46(24) |
| 43 | 17233992 | *rs8079562* | intron variant | G | A=0.25 | 3.30E-25 | 0.24(331) | -0.35(160) | -0.46(24) |
| 44 | 17234845 | *rs8065774* | intron variant | C | T=0.24 | 3.30E-25 | 0.24(331) | -0.35(160) | -0.46(24) |
| 45 | 17235918 | *rs41337846* | intron variant | T | C=0.24 | 6.60E-25 | 0.21(354) | -0.35(142) | -0.42(19) |

| S.No. | COORD | ID | HGVS | REF | ALT - FREQ (dbSNP) | SKIN (exposed) genotypes (normalized expression median values) | | | |
|---|---|---|---|---|---|---|---|---|---|
| | | | | | | **p-value** | **REF** | **HET** | **ALT** |
| 1 | 17236983 | *rs1708629* | 5' UTR variant | G | A=0.41 | 1.80E-55 | -0.38(162) | 0.01(263) | 0.27(180) |
| 2 | 17236986 | *rs41345949* | 5' UTR variant | C | T=0.05 | - | - | - | - |
| 3 | 17237171 | *rs1736209* | 5' UTR variant | C | G=0.72 | 1.00E-11 | 0.06(33) | -0.17(233) | 0.14(339) |
| 4 | 17212252 | *rs7218795* | 3' UTR variant | A | G=0.66 | 1.70E-20 | -0.3(44) | -0.19(234) | 0.19(327) |

| | | | | | | | | | |
|---|---|---|---|---|---|---|---|---|---|
| 5 | 17212319 | *rs7218992* | 3' UTR variant | C | A=0.06 | 4.30E-07 | 0.05(456) | -0.22(137) | -0.500(12) |
| 6 | 17213098 | *rs3803761* | 3' UTR variant | A | G=0.65 | 7.30E-20 | -0.20(46) | -0.19(235) | 0.19(324) |
| 7 | 17213262 | *rs12602675* | 3' UTR variant | C | T=0.08 | 1.10E-09 | 0.02(564) | -0.52(40) | N.A |
| 8 | 17216890 | *rs7208065* | intron variant | T | C=0.48 | 1.70E-54 | -0.62(129) | 0.03(296) | 0.30(180) |
| 9 | 17217030 | *rs41424546* | intron variant | C | T=0.108 | 1.30E-10 | 0.08(466) | -0.29(126) | -1.11(13) |
| 10 | 17217354 | *rs4985705* | intron variant | G | C=0.47 | 1.40E-57 | -0.63(130) | 0.03(292) | 0.29(183) |
| 11 | 17217498 | *rs4985752* | intron variant | T | C= 0.41 | 4.10E-56 | -0.46(161) | 0.04(268) | 0.28(176) |
| 12 | 17218310 | *rs8080386* | intron variant | T | C=0.60 | 1.70E-30 | -0.55(85) | -0.09(302) | 0.28(218) |
| 13 | 17219013 | *rs8065832* | intron variant | G | A=0.43 | 4.30E-45 | -0.48(162) | 0.04(294) | 0.28(149) |
| 14 | 17219819 | *rs2018781* | intron variant | G | C=0.73 | 9.50E-14 | -0.06(26) | -0.23(226) | 0.17(353) |
| 15 | 17221501 | *rs3744124* | intron variant | C | T=0.12 | 3.40E-18 | 0.06(529) | -0.44(76) | N.A |
| 16 | 17222119 | *rs1708622* | intron variant | A | G=0.74 | 1.80E-13 | -0.09(23) | -0.21(222) | 0.155(360) |
| 17 | 17222281 | *rs1736221* | intron variant | A | G=0.8 | 1.70E-30 | -0.55(85) | -0.09(302) | 0.28(218) |
| 18 | 17226610 | *rs1708620* | intron variant | C | T=0.43 | 9.60E-45 | -0.47(161) | 0.03(292) | 0.28(152) |
| 19 | 17226729 | *rs1708619* | intron variant | C | T= 0.44 | 6.70E-45 | -0.47(160) | 0.03(292) | 0.28(153) |
| 20 | 17226923 | *rs1736216* | intron variant | G | A=0.70 | 6.60E-08 | -0.03(44) | -0.15(242) | 0.13(319) |
| 21 | 17226931 | *rs6502565* | intron variant | C | T=0.13 | 1.30E-17 | 0.06(528) | -0.46(76) | N.A |
| 22 | 17226956 | *rs76319098* | intron variant | C | T=0.13 | 1.20E-17 | 0.05(548) | -0.55(56) | N.A |
| 23 | 17228555 | *rs1708618* | intron variant | T | C=0.47 | 2.50E-63 | -0.65(135) | 0.05(291) | 0.31(179) |
| 24 | 17228852 | *rs79717038* | intron variant | G | A=0.13 | 3.10E-19 | 0.06(543) | -0.58(61) | N.A |
| 25 | 17228888 | *rs1736215* | intron variant | G | A=0.46 | 5.30E-57 | -0.63(128) | 0.01(291) | 0.28(186) |
| 26 | 17229019 | *rs1736214* | intron variant | C | G=0.46 | 4.00E-56 | -0.59(126) | 0.01(290) | 0.29(189) |
| 27 | 17229304 | *rs1708617* | intron variant | A | G=0.72 | 7.70E-12 | 0.08(32) | -0.19(233) | 0.14(340) |
| 28 | 17229807 | *rs1613416* | intron variant | G | A=0.47 | 1.30E-56 | -0.62(129) | 0.01(288) | 0.28(188) |
| 29 | 17231214 | *rs1736213* | intron variant | T | G=0.48 | 5.50E-58 | -0.63(128) | 0.016(289) | 0.28(188) |
| 30 | 17231694 | *rs1736212* | intron variant | G | C=0.72 | 7.70E-12 | 0.08(32) | -0.19(233) | 0.14(340) |
| 31 | 17232108 | *rs1736211* | intron variant | A | G=0.74 | 8.40E-13 | -0.04(25) | -0.19(214) | 0.14(366) |
| 32 | 17233301 | *rs2349865* | intron variant | A | T=0.25 | 5.10E-12 | 0.08(33) | -0.19(233) | 0.14 (339) |
| 33 | 17233379 | *rs11078378* | intron variant | G | C=0.74 | 8.40E-13 | -0.04(25) | -0.19(214) | 0.14(366) |
| 34 | 17214314 | *rs8067893* | intron variant | G | A=0.11 | 1.70E-07 | 0.08(407) | -0.23(168) | 0.03 (30) |

| | | | | | | | | | |
|---|---|---|---|---|---|---|---|---|---|
| 35 | 17220853 | *rs41323249* | intron variant | C | T=0.12 | 4.40E-08 | 0.07(430) | -0.12(153) | -0.73(22) |
| 36 | 17221311 | *rs41400246* | intron variant | C | T=0.10 | 3.00E-10 | 0.07(430) | -0.25(129) | -1.11(13) |
| 37 | 17226117 | *rs41525346* | intron variant | A | G=0.08 | 8.70E-07 | 0.06(512) | -0.37(88) | N.A |
| 38 | 17227704 | *rs55836267* | intron variant | T | G= 0.12 | 4.40E-08 | 0.07(430) | -0.12(153) | -0.73(22) |
| 39 | 17230965 | *rs12602831* | intron variant | G | T=0.25 | 4.10E-27 | 0.13(419) | -0.33(163) | -1.05(23) |
| 40 | 17231193 | *rs12602871* | intron variant | G | A=0.25 | 4.10E-27 | 0.13(419) | -0.33(163) | -1.05(23) |
| 41 | 17233101 | *rs76045368* | intron variant | C | T=0.25 | 4.60E-27 | 0.15(390) | -0.27(184) | -0.83(31) |
| 42 | 17233347 | *rs34518797* | intron variant | C | T=0.25 | 4.60E-27 | 0.15(390) | -0.27(184) | -0.83(31) |
| 43 | 17233992 | *rs8079562* | intron variant | G | A=0.25 | 4.50E-27 | 0.15(391) | -0.25(183) | -0.83(31) |
| 44 | 17234845 | *rs8065774* | intron variant | C | T=0.24 | 4.50E-27 | 0.15(391) | -0.25(183) | -0.83(31) |
| 45 | 17235918 | *rs41337846* | intron variant | T | C=0.24 | 5.10E-28 | 0.14(417) | -0.33(164) | -1.06(24) |

**Note:** All these SNPs were common with our dataset

**Supplemental Table S10b: Alternate allele frequencies of non-coding variants of *FLCN* (found in our dataset) which are ≤ 15% in South Asian (gnomAD) and Indian (GenomeAsia100K) populations**

| S. No. | COORD | ID | HGVS | ANNOTATION | REF | ALT | gnomAD (South Asian) | Genome Asia100K (India) |
|---|---|---|---|---|---|---|---|---|
| 1 | 17212251 | *rs1451192210* | *c.*1404C>T* | 3' UTR | *G* | *T* | no pop freq | no pop freq |
| 2 | 17212319 | *rs7218992* | *c.*1336G>A* | 3' UTR | *C* | *A* | 0.06 | no pop freq |
| 3 | 17213230 | *rs7224335* | *c.*425G>A* | 3' UTR | *C* | *T* | 0.05 | 0.07 |
| 4 | 17213262 | *rs12602675* | *c.*393G>A* | 3' UTR | *C* | *T* | 0.08 | 0.08 |
| 5 | 17213299 | *rs7224474* | *c.*356G>T* | 3' UTR | *C* | *A* | 0.05 | 0.07 |
| 6 | 17214195 | *rs8068606* | *c.1539-339A>C* | intron | *T* | *G* | 0.04 | 0.06 |
| 7 | 17214314 | *rs8067893* | *c.1539-458C>T* | intron | *G* | *A* | 0.11 | no pop freq |
| 8 | 17214401 | *no ID* | *c.1539-545G>A* | intron | *C* | - | no pop freq | no pop freq |
| 9 | 17215375 | *rs34311146* | *c.1301-59C>T* | intron | *G* | *A* | 0.08 | 0.03 |
| 10 | 17215386 | *rs544930629* | *c.1301-70C>T* | intron | *G* | *T* | no pop freq | no pop freq |
| 11 | 17215879 | *rs572265105* | *c.1300+501A>G* | intron | *T* | *C* | 0.001 | 0.0008 |
| 12 | 17219825 | *rs564603245* | *c.872-616C>T* | intron | *G* | *T* | no pop freq | no pop freq |
| 13 | 17220853 | *rs41323249* | *c.871+684G>A* | intron | *C* | *T* | 0.12 | 0.09 |
| 14 | 17221311 | *rs41400246* | *c.871+226G>A* | intron | *C* | *T* | 0.1 | 0.05 |
| 15 | 17221501 | *rs3744124* | *c.871+36G>A* | intron | *C* | *T* | 0.12 | 0.11 |
| 16 | 17222727 | *rs2292527* | *c.619-66C>T* | intron | *G* | *A* | 0.17 | 0.13 |
| 17 | 17226117 | *rs41525346* | *c.396+59T>C* | intron | *A* | *G* | 0.08 | 0.04 |
| 18 | 17226931 | *rs6502565* | *c.250-609G>A* | intron | *C* | *T* | 0.13 | 0.12 |
| 19 | 17226956 | *rs76319098* | *c.250-634G>A* | intron | *C* | *T* | 0.13 | 0.12 |
| 20 | 17227493 | *rs373903620* | *c.249+396G>A* | intron | *C* | *T* | 0.015 | 0.009 |
| 21 | 17227704 | *rs55836267* | *c.249+185A>C* | intron | *T* | *G* | 0.12 | 0.09 |
| 22 | 17228404 | *rs75336342* | *c.-24-243G>A* | intron | *C* | *T* | 0.005 | 0.003 |
| 23 | 17228589 | *rs538804082* | *c.-24-428G>T* | intron | *C* | *A* | 0.17 | 0.13 |
| 24 | 17228852 | *rs79717038* | *c.-24-691C>T* | intron | *G* | *A* | 0.13 | 0.12 |
| 25 | 17229099 | *rs565915341* | *c.-24-938C>T* | intron | *G* | *A* | 0.005 | 0.009 |
| 26 | 17234361 | *rs375429199* | *c.-227-1460C>T* | intron | *G* | *A* | 0.04 | no pop freq |
| 27 | 17236986 | *rs41345949* | *c.-302G>A* | 5' UTR | *C* | *T* | no pop freq | 0.03 |

**Supplemental Table S11a: Pedigree Disequilibrium Test (PDT) for pathogenic *FLCN* mutations: pedigree informative parent triads ($X_T$) and discordant siblines ($X_S$) calculation to define random variable ($D$)**

| Family ID | Informative Parent ($X_T$) | Discordant sibships ($X_S$) | *D* |
|---|---|---|---|
| F1 | 2 ($n_t$ = 2) | 0 ($n_s$ = 1) | 0.66 |
| F2 | 0 ($n_t$ = 0) | -1 ($n_s$ = 1) | -1 |
| F4 | 0 ($n_t$ = 0) | 1 ($n_s$ = 1) | 1 |
| F5 | 3 ($n_t$ = 3) | 0 ($n_s$ = 0) | 1 |
| F11 | 1 ($n_t$ = 1) | 0 ($n_s$ = 1) | 1 |
| F12 | 2 ($n_t$ = 2) | 0 ($n_s$ = 1) | 1 |
| F13 | 0 ($n_t$ = 0) | 4 ($n_s$ = 4) | 4 |
| F15 | 1 ($n_t$ = 1) | 1 ($n_s$ = 1) | 1 |

**Supplemental Table S11b: Pedigree Disequilibrium Test (PDT) for SNP *rs1708629*: Pedigree triads ($X_T$) and siblines ($X_S$) calculation to define random variable ($D$)**

| Family ID | Informative Parent ($X_T$) | Discordant sibships ($X_S$) | *D* |
|---|---|---|---|
| F2 | 0 ($n_t$ = 0) | -2 ($n_s$ = 1) | -2 |
| F4 | 0 ($n_t$ = 0) | 0 ($n_s$ = 1) | 1 |
| F7 | 8 ($n_t$ = 4) | 0 ($n_s$ = 0) | 2 |
| F8 | 1 ($n_t$ = 1) | 1 ($n_s$ = 1) | 1 |
| F13 | 0 ($n_t$ = 0) | 8 ($n_s$ = 1) | 8 |
| F15 | 1 ($n_t$ = 1) | -5 ($n_s$ = 1) | -2 |

**Supplemental Table S11c: Pedigree Disequilibrium Test (PDT) for SNP *rs41345949*: Pedigree triads ($X_T$) and siblines ($X_S$) calculation to define random variable ($D$)**

| Family ID | Informative Parent ($X_T$) | Discordant sibships ($X_S$) | *D* |
|---|---|---|---|
| F4 | 0 ($n_t$ = 0) | 1 ($n_s$ = 1) | 1 |
| F6 | 0 ($n_t$ = 0) | 0 ($n_s$ = 1) | 0 |
| F7 | 8 ($n_t$ = 4) | 0 ($n_s$ = 0) | 2 |
| F8 | 0 ($n_t$ = 1) | -1 ($n_s$ = 1) | -0.5 |
| F15 | 2 ($n_t$ = 1) | -3 ($n_s$ = 1) | -0.5 |

**Supplemental Table S12: Effect of *FLCN* exonic mutations on protein structure**

| Effect of mutation | Exon 7 mutation | Exon 10 mutation | Exon 11 mutation | Exon 12 mutation |
|---|---|---|---|---|
| **Chromosomal position** | g.17222646 | g.17217085-17217095 | g.17216395 | g.17215284 |
| **HGVS** | *c.634C>T* | *c.1150_1160del delGTCCAGTCAGC* | *c.1285delC* | *c.1329_1332dupAGCC* |
| **Amino acid (AA) changes** | *Q212** | *p.V384F*2* | *p.H429T*39* | *p.A445S*11* |
| **Annotation** | Stop-gain | Frameshift, deletion | Frameshift, deletion | Frameshift, duplication |
| **Reference AA** | Glutamine | Phenylalanine | Histidine | Alanine |
| **Mutated AA** | Stop-codon | Valine | Threonine | Serine |
| **Protein Domain** | uDENN (N-terminal) | cDENN (C-terminal) | cDENN (C-terminal) | cDENN (C-terminal) |
| **CADD Score** | 43 (pathogenic) | 33 (pathogenic) | 33 (pathogenic) | 17.67 (pathogenic) |
| **Distance from Splice Site** | 16 | 17 | 16 | 33 |
| **position of stop codon in wt/ mu CDS** | 1740/636 | 1740/1155 | 1740/1401 | 1740/1368 |
| **position of stopcodon from site of mutation** | same | 2 | 39 | 11 |
| **Stop Codon wt/mu** | 580/212 | 580/385 | 580/467 | 580/457 |
| **Loss of protein features** | 11 helices, 5 beta strands, 1 turn, coiled-coil domain | 9 helices, 4 beta strands, 1 turn | 6 helives, 2 beta strands | 5 helives, 1 beta strand |

wt: wild-type, mu: mutated

**Note:** The N-terminal or longin terminal is part of upstream/uDENN domain (*p.86* to *p.242*), C-terminal is part of two domains - core/cDENN domain (*p.339* to *p.491*) and downstream/dDENN domain (*p.493* to *p.558*). For CADD, cut-off score of pathogenicity was decided to be 15, above which will be predicted to be pathogenic.

**Supplemental Table S13: Homology modelled structural validation of protein by PROCHECK (Ramachandran plot parameters)**

| Protein Structure (SWISSMODEL) | Ramachandran plot parameters (stereochemical properties) | | | |
|---|---|---|---|---|
| | Residues in core region | Residues in allowed region | Residues in disallowed region | Overall G-factor |
| FLCN (wild-type) | 80% | 17% | 0.20% | -0.28 |
| FLCN (ex7-*Q212**) | 80% | 17% | 0.60% | -0.27 |
| FLCN (ex10-*p.V384F*2*) | 79.90% | 17% | 0.30% | -0.3 |
| FLCN (ex11-*p.H429T*39*) | 82.80% | 14% | 1.10% | -0.24 |
| FLCN (ex12-*p.A445S*11*) | 86% | 11% | 0.50% | -0.27 |
| FNIP2 (wild-type) | 65% | 23% | 3.70% | -0.59 |
| RRAGA (wild-type) | 87.50% | 12.10% | 0.30% | -0.23 |
| RRAGC (wild-type) | 86% | 13.60% | 0% | -0.21 |

**Note:** PROCHECK analyses the stereochemical properties of protein structure by various parameters, with one of the key factors to be the Ramachandran plot. Residues are considered acceptable if more than 90% of the residues are in the core and overall allowed region. The G-factor measures the overall geometry, lower the G-factor, higher is a chance of something amiss in the geometry.

**Supplemental Table S14a: HADDOCK Scores for wild type or four-exonic mutant FLCN docked with wild-type FNIP2, RRAGA, and RRAGC (4-protein complex)**

| wt/mutant FLCN with wt FNIP2, RRAGA & RRAGC | *FLCN* mutations | HADDOCK score | Cluster size | RMSD from the overall lowest-energy structure | Van der Waals energy | Electrostatic energy | De-solvation energy | Restraints violation energy | Buried Surface Area | Z-Score |
|---|---|---|---|---|---|---|---|---|---|---|
| wt-FLCN | not applicable | -342.2±0.0 | 1 | 0.0±0.0 | -205.8±0.0 | -1036.0±0.0 | -30.1±0.0 | 1008.6±0.0 | 8467.7±0.0 | -2.1 |
| exon-7 mutant FLCN | *p.Q212* (c.634C>T)* | -203.2±30.5 | 2 | 25.2±3.0 | -150.7±7.3 | -746.8±39.4 | 2.5±3.2 | 944.3±120.6 | 5634.2±192.8 | 0 |
| exon-11 mutant FLCN | *p.V384F*2 (c.1150_1160 del11)* | -252.1±20.6 | 9 | 37.4±3.0 | -166.8±18.1 | -930.1±151.8 | -2.5±8.0 | 1032.5±70.0 | 6639.1±331.1 | 0 |
| exon-10 mutant FLCN | *p.H429T*39 (c.1285delC)* | -264.6±18.1 | 10 | 35.6±6.0 | -178.9±17.0 | -808.9±48.8 | -14.1±8.8 | 902.7±150.5 | 6172.7±268.3 | 0 |
| exon-12 mutant FLCN | *p.A445S*11 (c.1329_1332 dupAGCC)* | -242.8±2.8 | 8 | 35.2±4.5 | -163.6±10.0 | -837.7±78.8 | -13.2±10.5 | 1015.4±120.9 | 6171.2±341.2 | 0 |

Abbreviations: wt: wild-type, nuc: nucleotide

**Note:** HADDOCK is a protein-protein docking tool that docks protein monomers. Solvated docking analysis with FCC generated clusters with their corresponding desolvation energies. Z-scores indicate similarity between different clusters generated. All mutant FLCN monomers docked with wt-FNIP2-RRAGA-RRAGC generated only one cluster each, hence the z-score was zero in those four cases. About 200 clusters were generated with greater similarity (z-score -2.1) for wt-FLCN docked with wt-FNIP2-RRAGA-RRAGC. Greater negative z-scores indicate greater similarity between the clusters. The best cluster was selected based on the HADDOCK score, their z-score, buried surface areas and RMSD values.

**Supplemental Table S14b: HADDOCK Scores for wild type or four-exonic mutant FLCN docked with wild-type FNIP2 (2-protein complex)**

| wt/mutant ***FLCN*** **with wt-FNIP2** | **Mutated FLCN** | **HADDOCK score** | **Cluster size** | **RMSD from the overall lowest-energy structure** | **Van der Waals energy** | **Electrostatic energy** | **Desolvati on energy** | **Restraints violation energy** | **Buried Surface Area** | **Z-Score** |
|---|---|---|---|---|---|---|---|---|---|---|
| wt-FLCN | not applicable | -53.1±10.3 | 5 | 19.7±0.2 | -65.1±15.6 | -372.6±33.4 | -1.3±10.4 | 877.4±193.2 | 3280.5±460.2 | -1.5 |
| exon-7 mutant FLCN | *p.Q212* (c.634C>T)* | -47.9±11.9 | 7 | 20.7±0.1 | -64.5±5.8 | -130.9±21.4 | -11.7±3.0 | 544.5±49.1 | 2030.1±139.4 | -1.3 |
| exon-11 mutant FLCN | *p.V384F*2 (c.1150_1160del11)* | -32.0±19.4 | 4 | 7.1±0.3 | -49.9±6.3 | -257.9±65.5 | 0.7±4.7 | 687.8±35.9 | 2000.7±153.5 | -0.8 |
| exon-10 mutant FLCN | *p.H429T*39 (c.1285delC)* | -37.6±11.5 | 4 | 30.0±0.5 | -49.5±9.1 | -321.4±52.2 | 7.2±3.5 | 690.3±15.2 | 2201.0±90.2 | -1.5 |
| exon-12 mutant FLCN | *p.A445S*11 (c.1329_1332dupA GCC)* | -40.2±7.4 | 6 | 27.6±0.0 | -70.8±1.5 | -262.1±7.4 | 2.7±1.3 | 802.5±53.9 | 2485.0±39.3 | -1.3 |

Abbreviations: wt: wild-type, nuc: nucleotide

**Note:** Solvated docking analysis with FCC generated clusters generate their corresponding desolvation energies. Docking experiments generated ≥ 7 clusters for each complex. *Z*-scores indicate the structural similarity with the other clusters in the docking, the more negative z-score, and the better is the cluster. The best cluster was selected based on the HADDOCK score, their z-score, buried surface areas and RMSD values.

**Supplemental Table S15: Analysis of *FLCN* copy number variation in patients, asymptomatic members and unrelated healthy controls using Exons 4, 8 and 13 Taqman copy number assay**

| **Test: Paired t-test for Patients and Asymptomatics** | | | |
|---|---|---|---|
| **Groups** | **Exon 4 (p-value)** | **Exon 8 (p-value)** | **Exon 13 (p-value)** |
| Patients vs. Asymptomatics | 0.96 | 0.7 | 0.521 |
| **Test: Mann-Whitney test (Exon 4), Unpaired t-test (Exon 8 and 13)** | | | |
| **Groups** | **Exon 4 (p-value)** | **Exon 8 (p-value)** | **Exon 13 (p-value)** |
| Patients vs. Unrelated Controls | 0.825 | **0.019** | 0.186 |
| Asymptomatics vs. Unrelated controls | 0.348 | **0.008** | 0.125 |

**Note**: p-values <0.05 are given in bold. The $2^{-\Delta ct}$ values of the unrelated controls for only exon 4 assay were not in normal distribution (Kolmogorov-Smirnov tests), therefore, non-parametric Mann-Whitney tests were performed for Patients vs unrelated Controls, and Asymptomatics vs unrelated controls. Patient and asymptomatic groups were in normal distribution for all three exon assays (4, 8 and 13), therefore, parametric paired t-tests were performed between patients and asymptomatic members. Parametric unpaired t-tests were performed for exon 8 and 13 assays for patients vs unrelated controls and asymptomatic members vs unrelated controls, as unrelated controls for both assays were in normal distribution.

**Supplemental Table S16a: Germline variants in 3 genes – *SERPINA1*, *MTHFR* and *CBS* in 5 families (F6 to F10) by Sanger sequencing and their pathogenicity**

| GENE | COORD | ID | HGVS | Consequence | Annotation | Ref | Alt | Clinvar | F6 | F7 | F8 | F9 | F10 |
|---|---|---|---|---|---|---|---|---|---|---|---|---|---|
| ***SERPINA1*** | 14:94382864 | *rs709932* | *c.374G>A* | *p.Arg125His* | missense | *C* | *T* | Benign | | | | | |
| | 14:94390577 | *rs8004738* | *c.-458C>T* | *c.-458C>T* | 5' UTR | *G* | *A* | - | | | | | |
| | 14:94390673 | *rs8008743* | *c.-458C>T* | *c.-554G>C* | 5' UTR | *C* | *G* | - | | | | | |
| ***MTHFR*** | 1: 11794839 | *rs2066462* | *c.1056C>T* | *p.Ser352=* | synonymous | *G* | *A* | Likely benign | | | | | |
| | 1:11790767 | *rs797015332* | *c.1884G>A* | *p.Leu628=* | synonymous | *C* | *T* | - | | | | | |
| | 1:11790870 | *rs2274976* | *c.1781G>A* | *p.Arg594Gln* | missense | *C* | *T* | Likely benign | | | | | |
| | 1:11794395 | *rs1181242383* | *c.1310T>A* | *p.Leu437His* | missense | *A* | *T* | - | | | | | |
| | 1:11794400 | *rs4846051* | *c.1305C>T* | *p.Phe435=* | synonymous | *G* | *A* | Benign | | | | | |
| | 1:11794475 | *rs748977409* | *c.1230C>T* | *p.Ser410=* | synonymous | *G* | *A* | - | | | | | |
| | 1:11796321 | *rs1801133* | *c.665C>T* | *p.Ala222Val* | missense | *G* | *A* | pathogenic | | | | | |
| | 1:11802981 | *rs138189536* | *c.136C>T* | *p.Arg46Trp* | missense | *G* | *A* | pathogenic | | | | | |
| | 1:11803000 | *rs2066470* | *c.117C>T* | *p.Pro39=* | synonymous | *G* | *T* | Likely benign | | | | | |
| ***CBS*** | 21:43060506 | *rs1801181* | *c.1080C>T* | *p.Ala360=* | synonymous | *G* | *A* | Benign | | | | | |
| | 21:43065240 | *rs234706* | *c.699C>T* | *p.Tyr233=* | synonymous | *G* | *A* | Benign | | | | | |

**Note:** Coloured boxes indicate presence of variant in that family, F6, F7, F8 etc. Yellow: missense/pathogenic variants, green: synonymous variants, brown: UTR variants Bold indicates pathogenic missense variants.

**Supplemental Table S16b: Germline exonic and UTR variants found in 3 genes – *COL3A1*, *TSC1* and *TSC2* in 11 families (F1 to F11) by Targeted Amplicon NGS and their pathogenicity**

| **Gene: *COL3A1* (Chromosome 2)** | | | | | | | | | | | | | | | | | |
|---|---|---|---|---|---|---|---|---|---|---|---|---|---|---|---|---|---|
| **COORD** | **ID** | **HGVS** | **Consequence & Annotation** | **Ref** | **Alt** | **Clinvar** | **F2** | **F3** | **F4** | **F5** | **F6** | **F7** | **F8** | **F9** | **F10** | **F1-1** | **F11-70** |
| 188997371 | *rs7579903* | *c.1851G>A* | *p.Gln617Gln* (synonymous) | *G* | *A* | benign | | | | | | | | | | | |
| 188998698 | *rs1801183* | *c.2002C>A* | *p.Pro668Thr* (missense) | *C* | *A* | conflicting pathogenicity | | | | | | | | | | | |
| 188999354 | *rs1800255* | *c.2092G>A* | *p.Ala698Thr* (missense) | *G* | *A* | benign | | | | | | | | | | | |
| 188999856 | *rs1801184* | *c.2244T>C* | *p.Gly748Gly* (synonymous) | *T* | *C* | benign | | | | | | | | | | | |
| **Gene: *TSC1* (Chromosome 9)** | | | | | | | | | | | | | | | | | |
| 132891798 | *rs11553763* | *c.*4437G>A* | *c.*4437G>A* (3' UTR) | *C* | *T* | Benign | | | | | | | | | | | |
| 132892247 | *rs563835484* | *c.*3988G>A* | *c.*3988G>A* (3' UTR) | *C* | *T* | Likely benign | | | | | | | | | | | |
| 132892556 | *rs1050700* | *c.*3679G>A* | *c.*3679G>A* (3' UTR) | *C* | *T* | Benign | | | | | | | | | | | |
| 132894301 | *rs10491534* | *c.*1934A>G* | *c.*1934A>G* (3' UTR) | *T* | *C* | Benign | | | | | | | | | | | |
| 132894728 | *rs739441* | *c.*1507G>A* | *c.*1507G>A* (3' UTR) | *C* | *T* | Benign | | | | | | | | | | | |
| 132894747 | *rs739442* | *c.*1488C>T* | *c.*1488C>T* (3' UTR) | *G* | *A* | Benign | | | | | | | | | | | |
| 132894863 | *rs58612431* | *c.*1372T>G* | *c.*1372T>G* (3' UTR) | *A* | *C* | Benign | | | | | | | | | | | |
| 132894913 | *rs2809243* | *c.*1322C>T* | *c.*1322C>T* (3' UTR) | *G* | *A* | Benign | | | | | | | | | | | |
| 132894960 | *rs2809244* | *c.*1275T>G* | *c.*1275T>G* (3' UTR) | *A* | *C* | Benign | | | | | | | | | | | |

| | | | | | | |
|---|---|---|---|---|---|---|
| 132895322 | *rs117425923* | *c.*913C>T* | *c.*913C>T* (3' UTR) | *G* | *A* | Likely benign |
| 132895394 | *rs149902841* | *c.*841C>T* | *c.*841C>T* (3' UTR) | *G* | *A* | Likely benign |
| 132895946 | *rs11323835* | *c.*289delT* | *c.*289delT* (3' UTR) | *A* | *A/-* | Benign |
| 132897330 | *rs4962081* | *c.2829C>T* | *p.Ala943Ala* (synonymous) | *G* | *A* | Benign |
| 132935081 | *rs116951280* | *c.-129A>T* | *c.-129A>T* (5' UTR) | *T* | *A* | Likely benign |
| **Gene: *TSC2* (Chromosome 16)** | | | | | | |
| 2056721 | *rs570051626* | *c.726C>T* | *p.Thr242Thr* (synonymous) | *C* | *T* | Likely benign |
| 2064406 | *rs34012042* | *c.1578C>T* | *p.Ser526Ser* (synonymous) | *C* | *T* | Likely benign |
| 2084437 | *rs45517331* | *c.4215C>T* | *p.Ala1405Ala* (synonymous) | *C* | *T* | Likely benign |
| 2084981 | *rs137854239* | *c.4527_4529 delCTT* | *p.Phe1510del* (synonymous) | *CT T* | *CT T/-* | Likely benign |
| 2088268 | *rs1748* | *c.5202T>C* | *p.Asp1734Asp* (synonymous) | *T* | *C* | Benign |
| 2088583 | *rs1051771* | *c.5397G>C* | *p.Ser1799Ser* (synonymous) | *G* | *C* | Likely benign |
| 2088670 | *rs36032671* | *c.*60_*61de lA* | *c.*61_*62del* (synonymous) | *AA* | *AA/ -* | uncertain significance |

**Note:** COORD: chromosomal position. Coloured boxes indicate presence of the variant in that family F2, F3, F4 etc. Missense variants are denoted by yellow, synonymous variants in green, deletions in grey and UTR variants in brown. Variants were found in both patients and asymptomatic members.

**Supplemental Table S17: Demography and presence of *FLCN* pathogenic mutations in 27 patients suffered from PSP recurrence**

| Family ID | Patient ID | Age-range for onset of 1st PSP | PSP occurrences (no. of times) | Sex | Family history | Smoking (≥10 years) | Pathogenic *FLCN* mutations |
|---|---|---|---|---|---|---|---|
| F1 | F1-1 | 31-35 | 1 | female | present | absent | present |
| | F1-2 | 26-30 | 1 | female | present | absent | present |
| F2 | F2-9 | 41-45 | 1 | male | absent | absent | present |
| F3 | F3-13 | 36-40 | 3 | male | present | absent | present |
| | F3-14 | 36-40 | 1 | male | present | absent | present |
| F4 | F4-18 | 41-45 | 2 | male | absent | absent | present |
| F5 | F5-25 | 31-35 | 1 | female | present | absent | present |
| | F5-26 | 26-30 | 1 | male | present | absent | present |
| | F5-28 | 21-25 | 2 | male | present | absent | present |
| F6 | F6-35 | 61-65 | 1 | male | absent | absent | absent |
| F7 | F7-44 | 15-20 | 4 | male | present | absent | absent |
| | F7-45 | 31-35 | 1 | female | present | absent | absent |
| | F7-46 | 31-35 | 1 | male | present | absent | absent |
| | F7-47 | 36-40 | 2 | female | present | absent | absent |
| | F7-48 | 55-60 | 2 | female | present | absent | absent |
| F8 | F8-56 | 15-20 | 1 | female | present | absent | absent |
| F10 | F10-67 | 56-60 | 1 | female | absent | present | absent |
| F11 | F11-70 | 21-25 | 4 | female | present | absent | present |
| F12 | F12-77 | 56-60 | 1 | female | present | absent | present |
| | F12-78 | 41-45 | 1 | male | present | present | present |
| F13 | F13-82 | 46-50 | 2 | male | present | present | present |
| | F13-83 | 46-50 | 1 | male | present | present | present |
| | F13-84 | 41-45 | 1 | male | present | present | present |
| | F13-85 | 31-35 | 2 | male | present | present | present |
| F14 | F14-95 | 31-35 | 1 | female | absent | absent | present |
| F15 | F15-99 | 51-55 | 1 | male | present | absent | present |
| | F15-101 | 31-35 | 1 | female | present | absent | present |

**Note:** GEE (SPSS) was used for analyzing the probability of a recurrence of a PSP with age. Since, families imply clustered data, hence GEE was used for this analysis. The number of times (PSP occurrences) was taken as a dependent variable for 'age of onset of first PSP', while patient sex, family history, smoking habits and presence of pathogenic *FLCN* mutations were taken as co-factors. PSP occurrences: 1 (PSP has occurred once), 2 (PSP has occurred twice), 3 (PSP has occurred thrice), 4 (PSP has occurred four times). Other factors were taken as binary (as presence or absence). Pleurodesis was not taken as co-factor, as none of the patients opted for the procedure after their first PSP. Size and number of lung cysts were not considered.

**Supplemental Table S18: Analysis of copy number variation for patients, asymptomatic members (families F3, F4, F10) and unrelated healthy controls using Taqman data of Exons 4, 8 and 13**

| **Test: Paired t-test for Patients and Asymptomatics** | | | |
|---|---|---|---|
| **Groups** | **Exon 4 (p-value)** | **Exon 8 (p-value)** | **Exon 13 (p-value)** |
| Patients vs. Asymptomatics | 0.719 | 0.261 | 0.744 |
| **Test: Mann-Whitney test (Exon 4), Unpaired t-test (Exon 8 and 13)** | | | |
| **Groups** | **Exon 4 (p-value)** | **Exon 8 (p-value)** | **Exon 13 (p-value)** |
| Patients vs. Controls | **0.049** | **0.037** | 0.313 |
| Asymptomatics vs. Controls | **0.027** | **0.027** | 0.188 |

**Note:** p-values <0.05 are shown in bold. Non-parametric Mann-Whitney tests were performed for Patients vs Controls and Asymptomatics vs controls only for Exon 4, as the $2^{-\Delta ct}$ values of the controls were not in normal distribution (Kolmogorov-Smirnov tests). Parametric paired t-tests were performed between patients and asymptomatic members for all 3 exon assays, while unpaired t-tests were performed for exon 8 and 13 assays for patients vs controls and asymptomatic members vs controls.
