## Supplemental Figures for "Genetic Insight into Birt-Hogg-Dubé syndrome in Indian patients reveals novel mutations in *FLCN*"

### Supplemental Figure S1: Patient phenotype ontology distribution

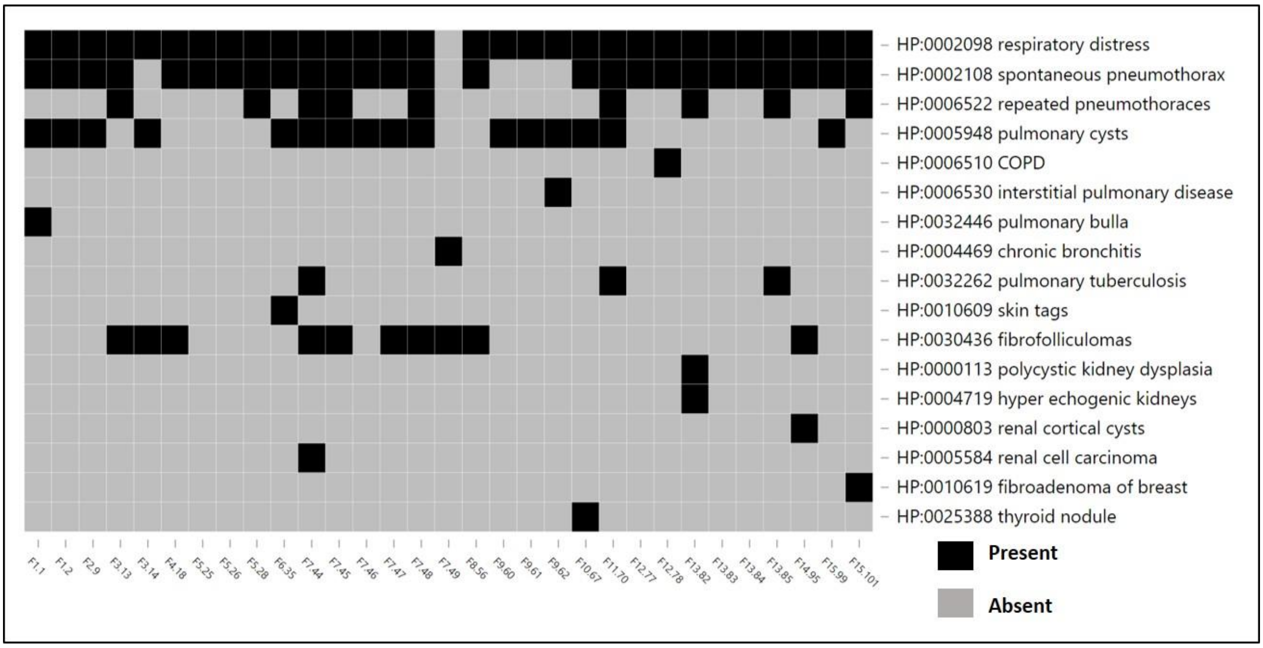


**Legend**: X-axis: anonymized patient IDs, Y-axis: presence/absence of HPO terms.

Black: presence, grey: absence of HPO terms

**Supplemental Figure S2: HPO-Phenomizer results for 31 patients**

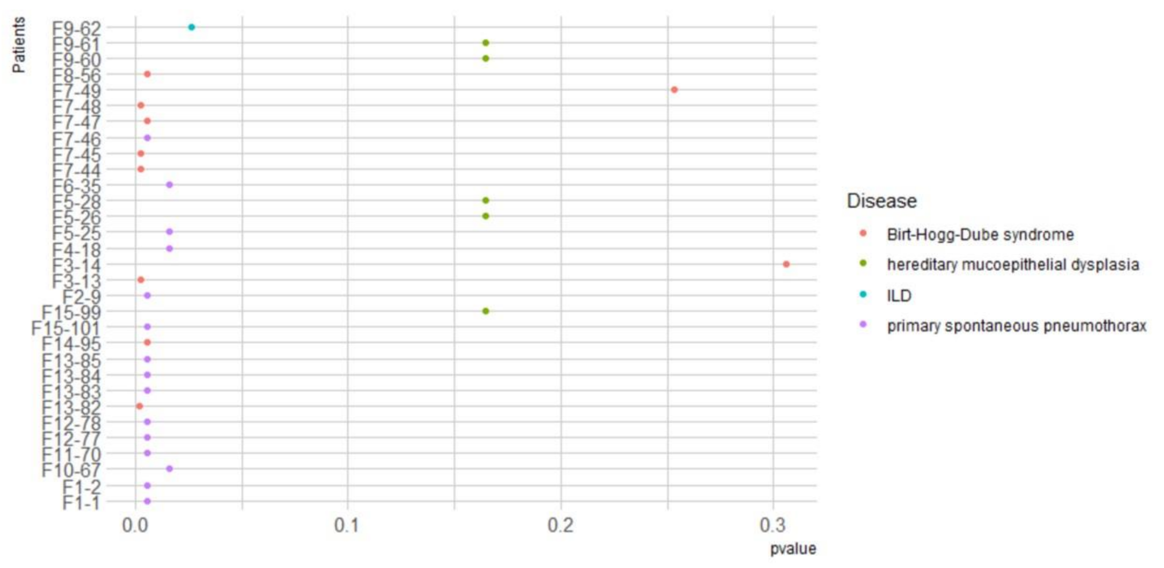


**Legend:** X-axis: p-values and Y-axis: anonymized patient IDs. ILD: Interstitial lung disease *P*-values of the ranked diseases for each patient was calculated by Benjamini-Hochberg multiple testing correction. Diseases are denoted by different coloured dots.. Phenomizer assesses the HPO terms for each patient and assigns a rank based on its p-value using semantic similarity search of various Mendelian diseases. Primary spontaneous pneumothorax (PSP) and Birt-Hogg-Dubé syndrome (BHDS) were found in 15 and 8 patients, respectively, with a significant p-value (≤ 0.05). Two patients were diagnosed with BHDS, but with a higher p-value (> 0.2).

**Supplemental Figure S3: Distribution of variants in different regions of *FLCN* in patients and asymptomatic family members taken for targeted amplicon NGS**

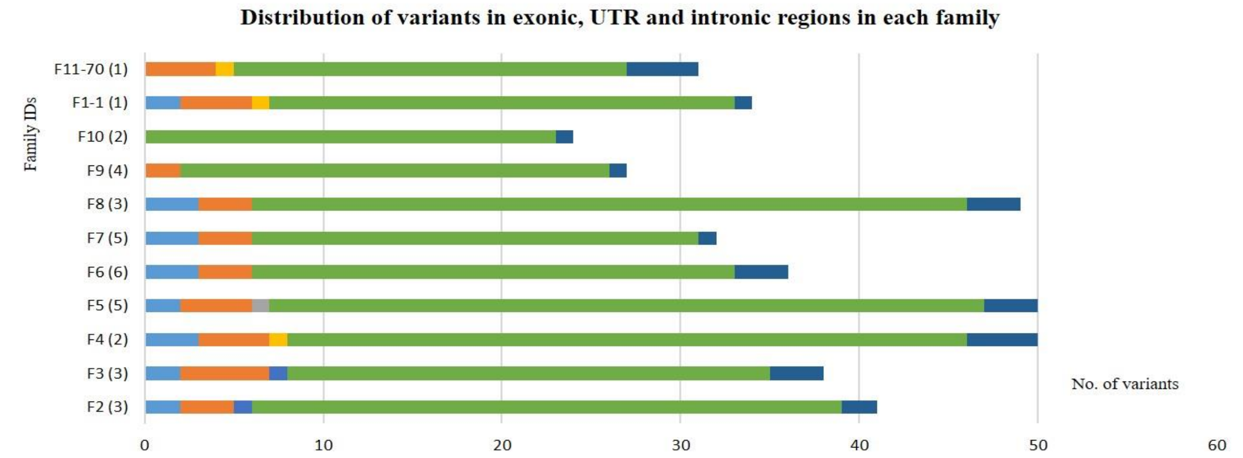


| | F2 (3) | F3 (3) | F4 (2) | F5 (5) | F6 (6) | F7 (5) | F8 (3) | F9 (4) | F10 (2) | F1-1 (1) | F11-70 (1) |
|---|---|---|---|---|---|---|---|---|---|---|---|
| 5' UTR | 2 | 2 | 3 | 2 | 3 | 3 | 3 | 0 | 0 | 2 | 0 |
| 3' UTR | 3 | 5 | 4 | 4 | 3 | 3 | 3 | 2 | 0 | 4 | 4 |
| Exonic SNV | 0 | 0 | 0 | 1 | 0 | 0 | 0 | 0 | 0 | 0 | 0 |
| Exonic Indels | 0 | 0 | 1 | 0 | 0 | 0 | 0 | 0 | 0 | 1 | 1 |
| Splicing | 1 | 1 | 0 | 0 | 0 | 0 | 0 | 0 | 0 | 0 | 0 |
| Intronic SNV | 33 | 27 | 38 | 40 | 27 | 25 | 40 | 24 | 23 | 26 | 22 |
| Intronic Indels | 2 | 3 | 4 | 3 | 3 | 1 | 3 | 1 | 1 | 1 | 4 |

5' UTR  3' UTR  Exonic SNV  Exonic Indels  Splicing  Intronic SNV  Intronic Indels

**Legend**: Colour codes and number of variants in each section are given in the table at the bottom of the chart. Number of variants in UTR, exonic and intronic regions in each family are mentioned. F1-1 and F11-70 are the only patients from family F1 and F11. Numbers in the bracket correspond to the number of individuals taken for NGS study from those families, eg. F11-70 (1): family number-F11; member ID-70; only 1 sample taken from family F11.

**Supplemental Figure S4: Pathogenic *FLCN* mutations: Chromatograms of pathogenic mutations in *FLCN***

**S4-1. Stop-gain mutation (*c.634C>T in exon 7*)**

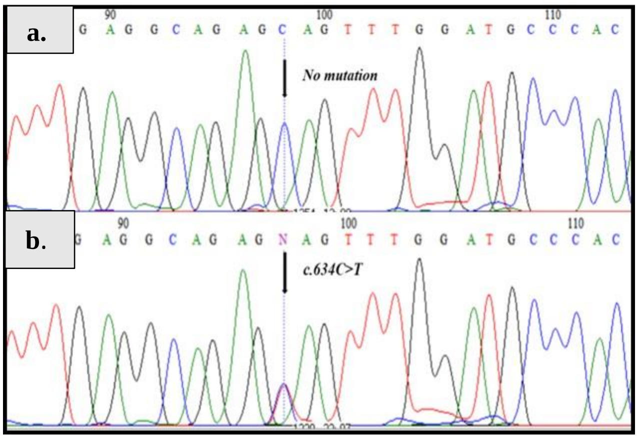


**S4-2. Eleven-nucleotide deletion mutation (*c.1150_1160delGTCCAGTCAGC in exon 10*)**

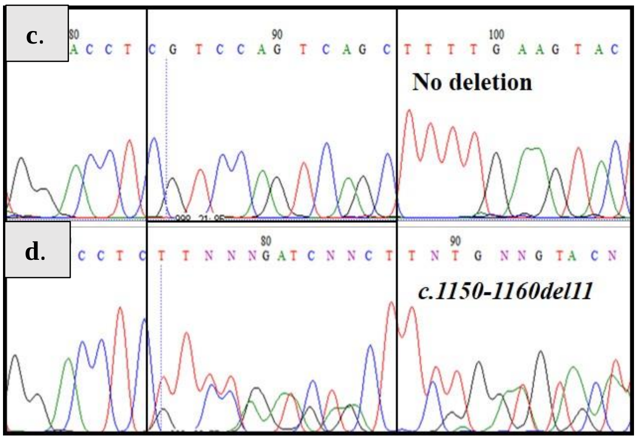


**S4-3. Hotspot single-nucleotide deletion mutation (*c.1285delC in exon 11*)**

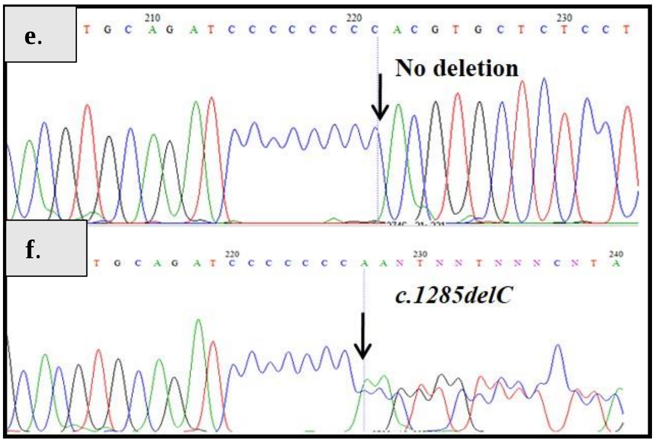

**S4-4. Four-nucleotide duplication mutation (*c.1329_1332dupAGCC in exon 12*)**

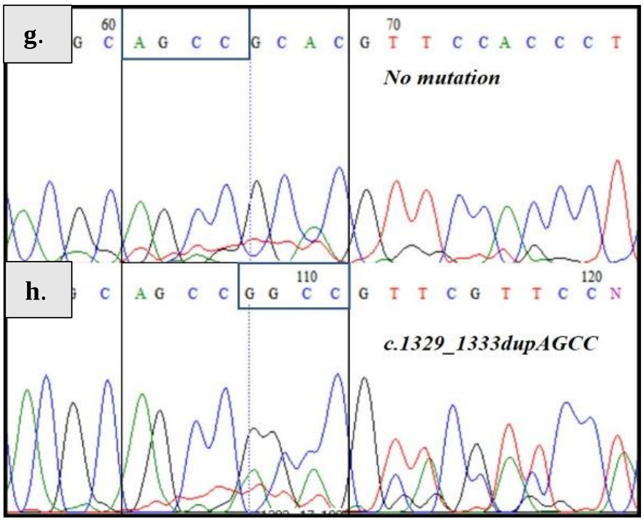


**S4-5. Splice donor mutation (*c.1300+1G>A* in exon-intron boundary of exon 11)**

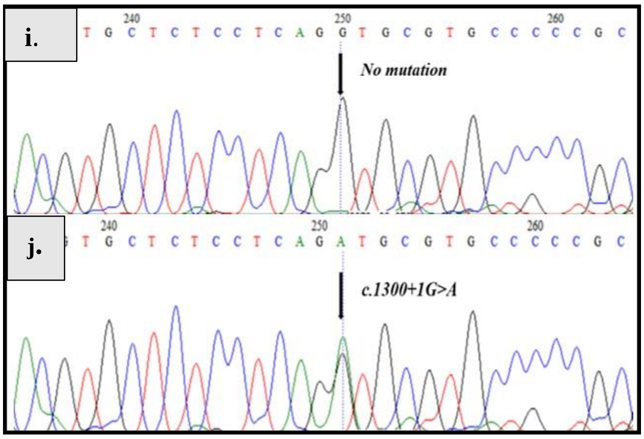


**S4-6. Splice acceptor mutation (*c.1301-1G>A* in exon-intron boundary of exon 12)**

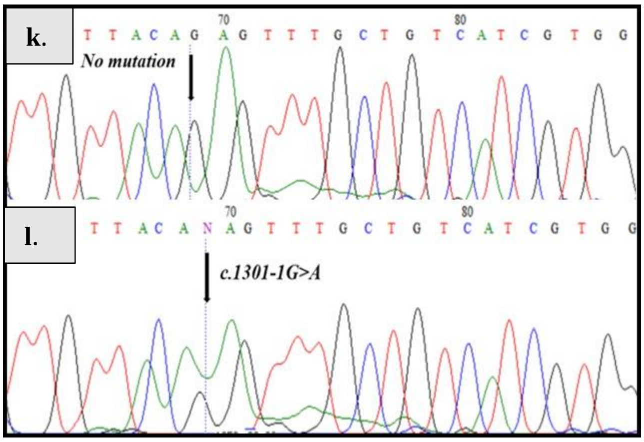

**Legend:** Chromatograms were obtained using Sanger sequencing method for both normal and mutated sequences.

a) Asymptomatic member F5-27 with no mutation and b) patient F5-25 with heterozygous *c.634C>T* stop-gain mutation;

c) Asymptomatic F11-75 member with no mutation and d) patient F11-70 with heterozygous deletion mutation *c.1150_1160del11*;

e) Asymptomatic member F12-79 with no mutation and f) patient F12-78 with heterozygous deletion *c.1285delC* mutation;

g) Asymptomatic member F4-19 with no mutation and h) patient F4-18 with heterozygous duplication *c.1329_1332dupAGCC* mutation;

i) Asymptomatic member F2-11 with no mutations and j) patient F2-9 with heterozygous splice donor variant *c.1300+1C>T* (splice region exon 11-12 boundary);

k) Asymptomatic member F3-15 with no mutation and l) patient F3-13 with heterozygous splice acceptor variant *c.1301-1C>T* ;

All mutations are heterozygous. So, chromatograms S4-2d, S4-3f, S4-4h are heterozygous deletions or duplication, where the non-mutated sequence slides over the deleted and duplicate regions, creating a noisy sequence thereafter the point of deletion/duplication as indicated. Heterozygous single nucleotide variations have two peaks of two alleles at the same locus (S4-1b, S4-5j, S4-6l). Arrows or rectangular boxes indicate regions of mutations.

**Supplemental Figure S5a: Protein domains of FLCN (SWISSMODEL)**
(Blue: N-terminal, brown: C-terminal)

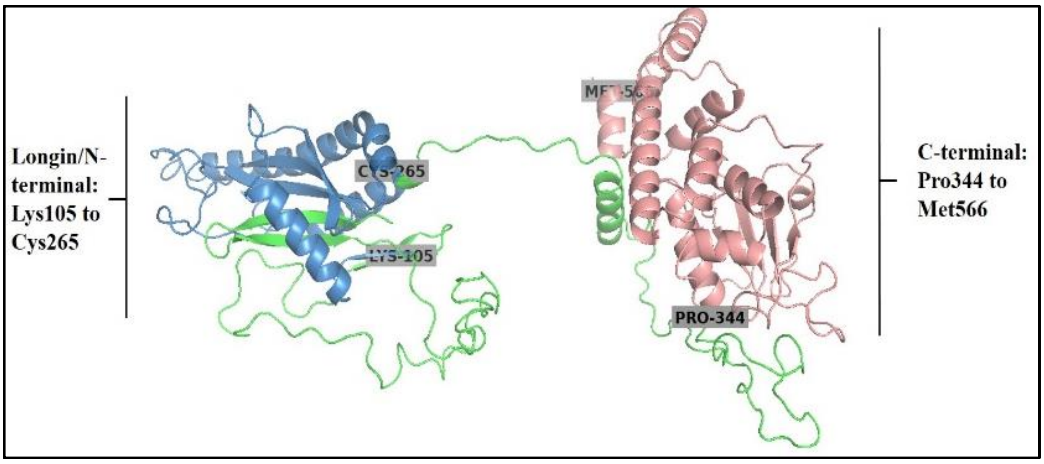


**Legend:** FLCN is homology modelled using SWISSMODEL based on the L-chain of pdb structure - *6ulg*. FLCN contains two terminals Longin/N-terminal (*Lys105* to *Cys265*) and C-terminal (*Pro344* to *Met566*). The two terminals are indicated by different colours, with their start and end residues.

**Supplemental Figure S5b: FLCN exonic mutations mapped to protein domains**

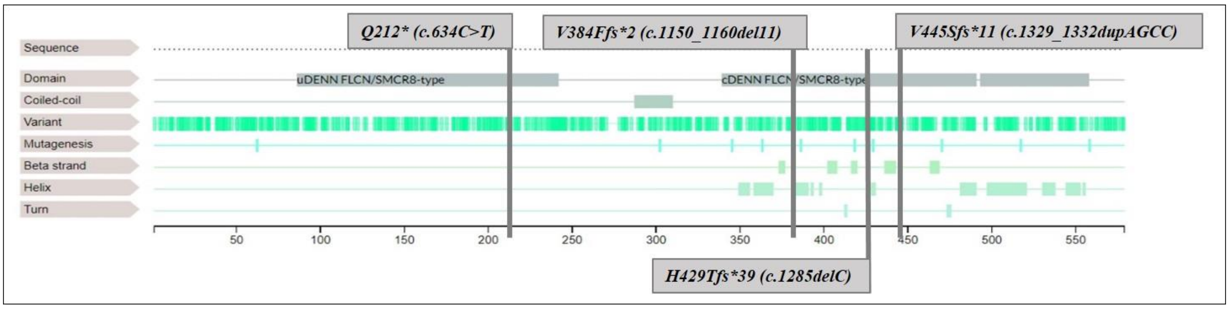


**Legend**: Linear representation of the FLCN sequence, domains, known mutations and variant site with secondary structure. Four pathogenic mutations of FLCN mapped to their corresponding position on the protein sequence.

**Supplemental Figure S6a: Four-protein docking structures of FLCN-FNIP2-RRAGA-RRAGC with: i) wild type (wt) FLCN ii) exon7-mutant-FLCN iii) exon10-mutant-FLCN iv) exon11-mutant-FLCN v) exon12-mutant-FLCN**

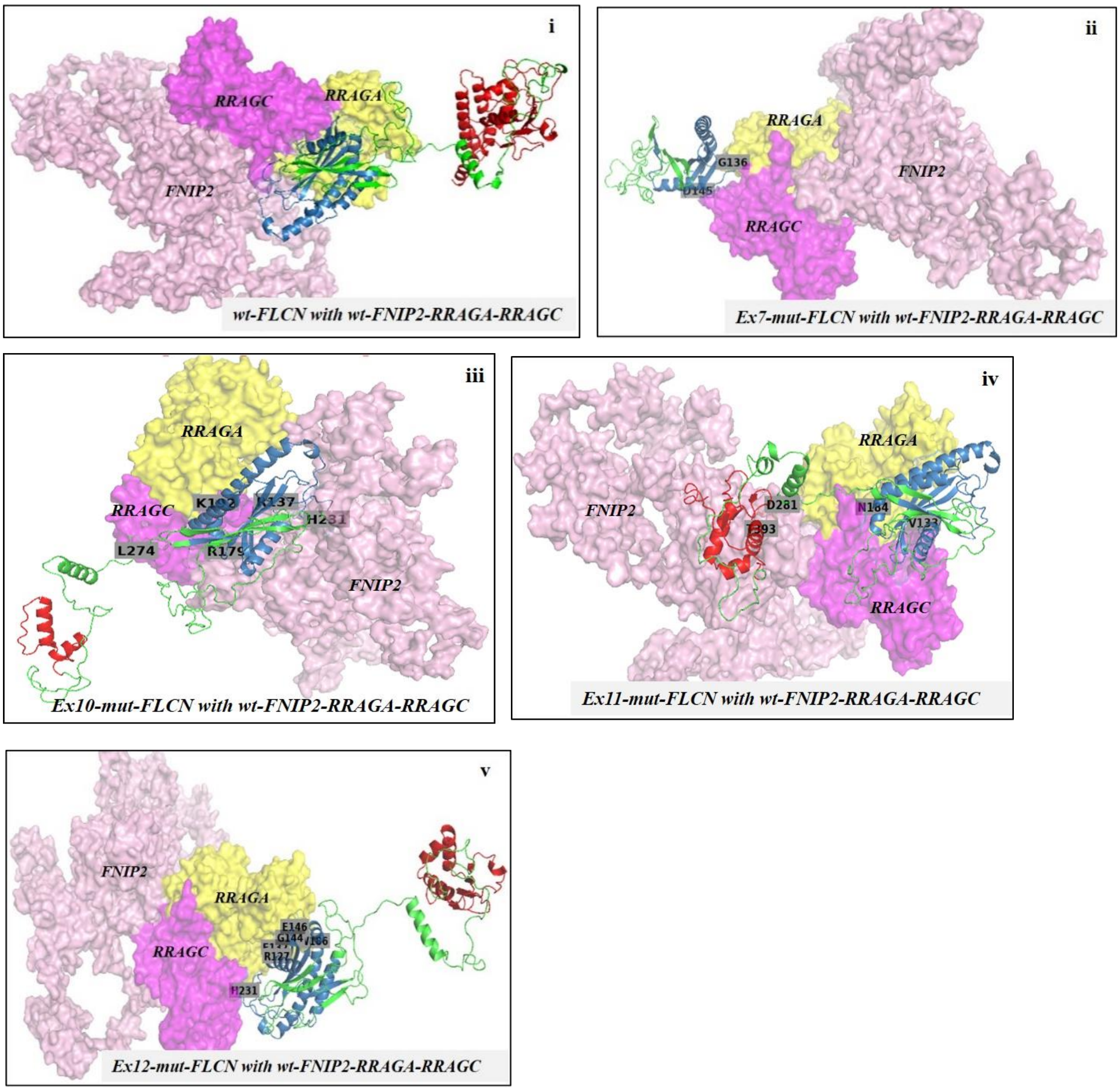


**Legend:** Pink surface: FNIP2, yellow: RRAGA, magenta: RRAGC. FLCN is depicted in green with N-terminal **in blue**, and C-terminal **in red**. Interactive residues of *FLCN* were extracted using a 5Å cut off and denoted by grey rectangles.

Wild-type RRAGA-RRAGC-FNIP2 docked with –

**i) wild-type FLCN**: The C-terminal of *FLCN* does not seem to interact with wild-type FNIP2-RRAGA-RRAGC complex in this docked complex. wt-FLCN interacts via *Glu132*, *Pro140*, *Glu146*, *Arg179* and *Lys77*, *Gln123*, *Arg137*, *Leu230*, *Gln232* (last 5 residues not found in wild-type FLCN interactive interface of *6ulg*).

**ii) exon7-mut-FLCN**: The stop-gain mutated and truncated FLCN sparsely interacts with RRAGA and RRAGC via *Asp145* and *Gly136*.

**iii) exon10-mut-FLCN**: The 11-nucleotide deletion and truncated FLCN interacts via *His231*, *Arg137, Arg 179*, *Lys192,* and *Leu274*. Last two residues were not found in the wild-type FLCN interactive interface (in *6ulg*).

**iv) exon11-mut-FLCN**: The 'hotspot-mutant' and truncated FLCN interacts via *Val133, Glu141, Asn184, Asp281* and *Thr393*. None of these residues are common with the interactive interfaces in *6ulg*.

**v) exon12-mut-FLCN**: The 4-nucleotide duplicated and truncated FLCN interacts via *His231*, *Glu132, Glu146, Arg127, Gly144,* and *Trp186*. The last three residues were not found in wild-type FLCN interactive interface (in *6ulg*).

**Supplemental Figure S6b: Two-protein docking model between wild type-FNIP2 and – i) wt FLCN ii) exon7-mutant-FLCN iii) exon10-mutant-FLCN iv) exon11-mutant-FLCN v) exon12-mutant-FLCN**

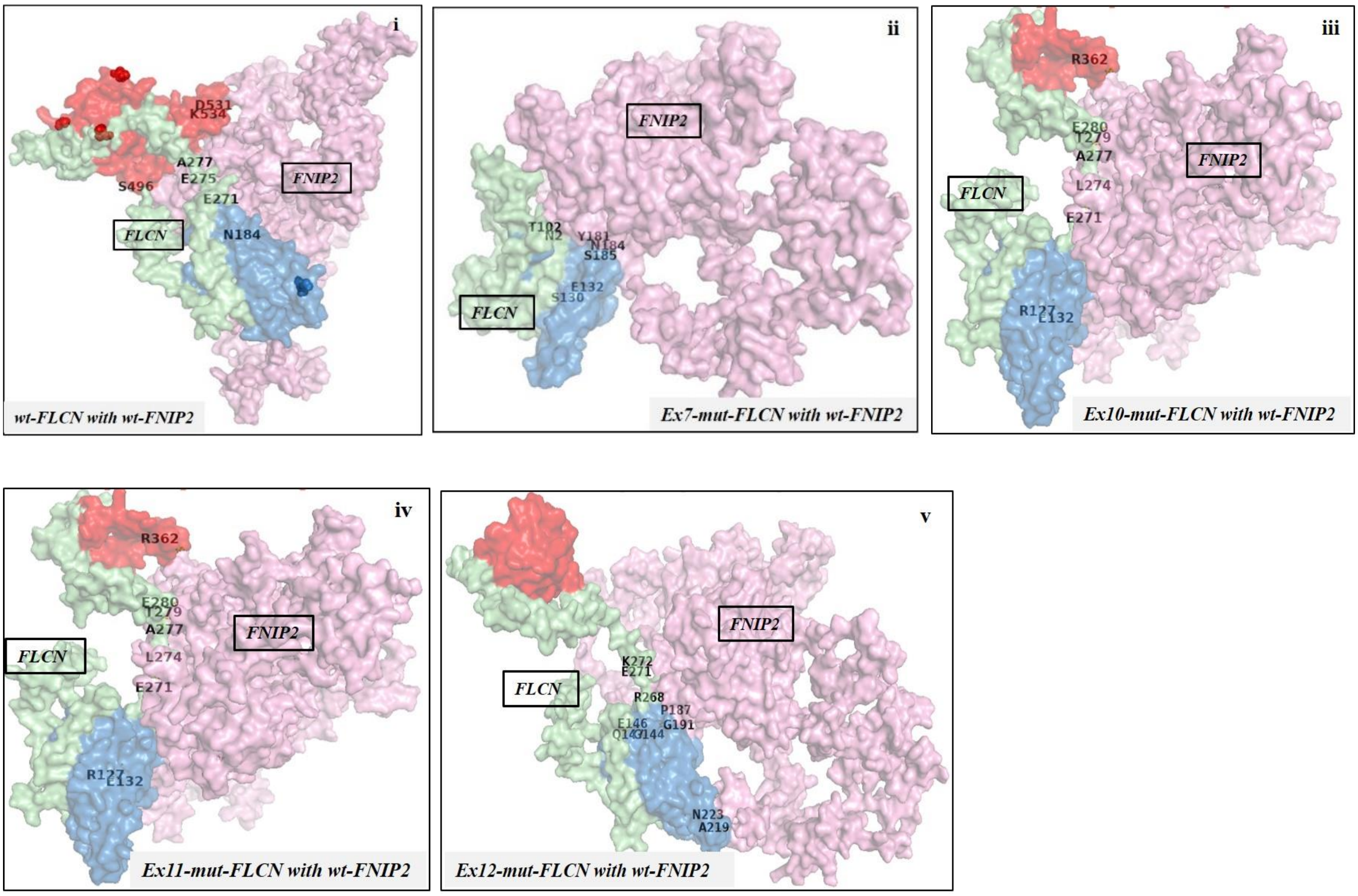


**Legend:** Pink: FNIP2, Green: FLCN with blue depicting N-terminal and red depicting C-terminal of protein. The sites of mutation - *Q212, V384, H429, A445* have been shown as spheres in Fig. i. Interactive residues of FLCN were extracted using a 5Å cut off and those residues have been shown.

Wild-type FNIP2 docked with –

**i) wild-type FLCN**: interacts via *Pro140*, *Ala277* and *Arg 577* (common in the interactive residues of FLCN in *6ulg*), along with *Asn184*, *Asn492*, *Asn494*, *Ala488*, *Ser526*.

**ii) exon7-mut-FLCN:** The stop-gain mutated and truncated FLCN interacts via *Asn2, Thr102, Ser130, Glu132, Tyr181, Asn184* and *Ser185.*

**iii) exon10-mut-FLCN:** The 11-nucleotide deletion and truncated FLCN interacts via *Arg127, Glu132, Glu271, Leu274, Ala277, Thr279, Glu280* and *Arg362.*

**iv) exon11-mut-FLCN:** The 'hotspot-mutant' and truncated FLCN interacts with FNIP2 via *Gln123, Arg179, Asn184, Glu271, Leu274, Gly276, Ala277* and *Met394. Asn184* is common with one of the interacting residues of
ex11-mut-FLCN with wt-FNIP2 in the 4-protein docked structure as well.

**v) exon12-mut-FLCN:** The 4-nucleotide duplicated and truncated FLCN interacts via *Gly144, Glu146, Gln147, Pro187, Gly191, Ala219, Asn223, Arg268, Glu271* and *Lys272*. *Glu146* is common with one of the interacting residues of ex12-mut-FLCN with wt-FNIP2 in the 4-protein docked complex.

**Supplemental Figure S7: Log transformed read counts in five regions of *FLCN* (analysed in Seqmonk) in 35 samples taken for targeted amplicon NGS**

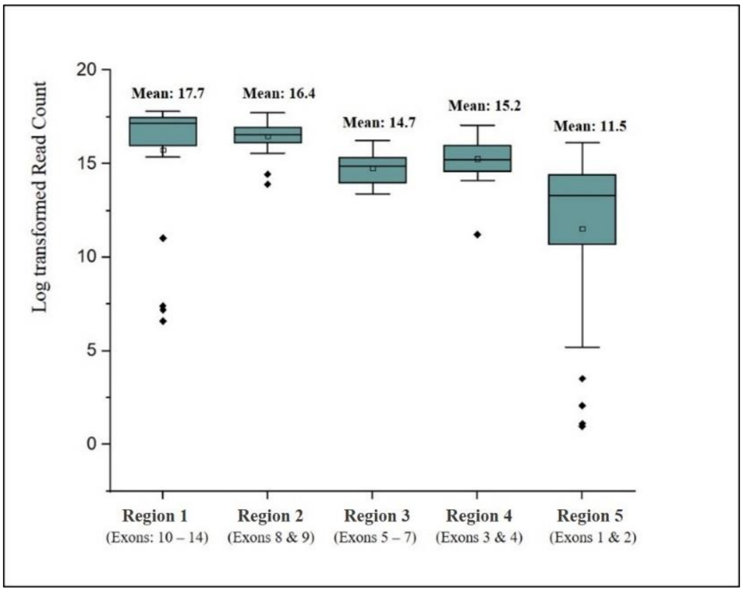


**Legend:** Regions 1 to 5 depict the five regions of *FLCN* divided for read-count normalization in Seqmonk. Region 1 encompasses exons 10, 11, 12, 13 & 14 of *FLCN*, region 2 encompasses exons 8 & 9, Region 3 includes exons 5, 6 & 7, Region 4 include exons 3 & 4, and Region 5 contains exons 1 & 2.  The boxplots represents the normalized log transformed read count of 20 patients and 15 asymptomatic members. Black diamonds outside the plots represent the outliers indicating a very low read count.

The samples representing the outliers in the samples for, Region 1:  F3-13 (patient, family F3), F9-61 (patient, family F9), F10-67 (patient, family F10), F10-68 (asymptomatic member, family F10); Region 2: F4-18 (patient, family F4), F5-23 (asymptomatic member, family F5); Region 3: F6-36 (asymptomatic member, family F6); and Region 5: F6-36 (asymptomatic member, family F6), F9-62 (patient, family F9), F10-67 (patient, family F10), F10-68 (asymptomatic member, family F10), F11-70 (patient, family F11).

Two asymptomatic members, F5-23 and F6-36 were not considered for further analysis owing to their low amplicon concentration after the initial PCR amplification. Read counts of the 5 outliers for Region 5 (encompassing exons 1 & 2) ranged from 0.9 to 3.5 with barely any read depth in the overall region. Hence, data of these 5 samples were not analysed further. Therefore, normalized read-count differences between patients and asymptomatic members particularly in Regions 1 and 2 of *FLCN* were observed in four families (F3, F4, F9 and F10).

**Supplemental Figure S8a and S8b: Chromatograms of *MTHFR* pathogenic SNPs**

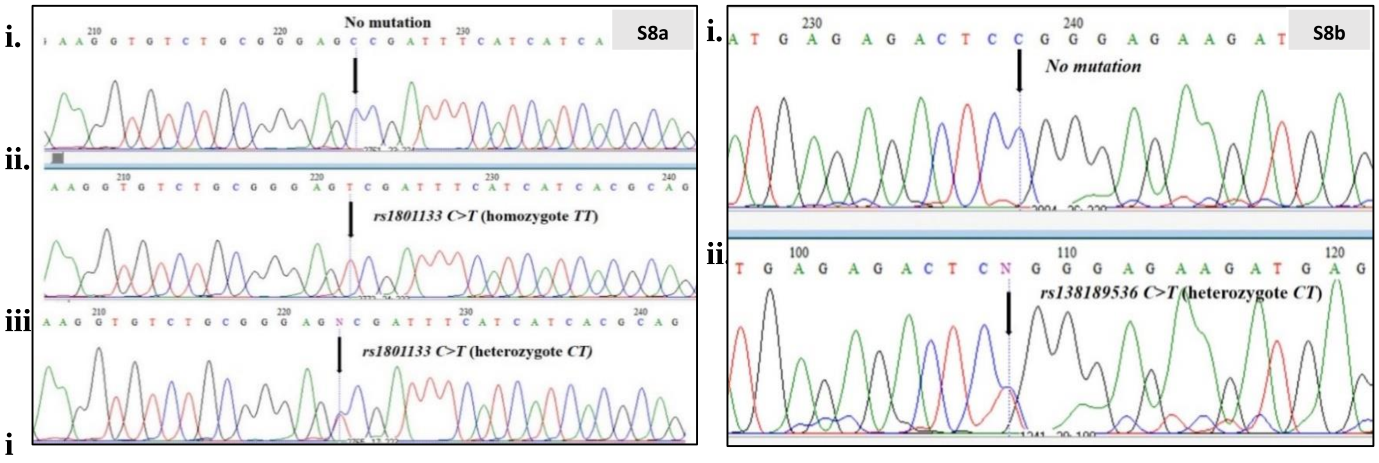


**Legend: S8a**. i) asymptomatic member (F9-66) with homozygote reference *CC,* (ii) parent of Index (F9-61) patient with homozygous alternate *TT* for *rs1801133* and iii) Index patient (F9-60) with heterozygote *CT*

**S8b.** Chromatograms of (i) asymptomatic member (F9-66) with homozygote reference *CC* (ii) Index patient (F9-60) with heterozygote *CT* for *rs138189536*
